# Tracking patient clusters over time enables to extract all the information available in the medico-administrative databases

**DOI:** 10.1101/2022.08.05.22278468

**Authors:** Judith Lambert, Anne-Louise Leutenegger, Anne-Sophie Jannot, Anaïs Baudot

## Abstract

**Context:** Identifying clusters (i.e., subgroups) of patients from the analysis of medico-administrative databases is particularly important to better understand disease heterogeneity. However, the complexity of these databases, in particular due to the presence of truncated longitudinal data, requires adaptation of clustering approaches.

**Objective:** We propose here cluster-tracking approaches to identify clusters of patients from longitudinal data contained in medico-administrative databases.

**Material and Methods:** We first cluster patients at each age using either the Markov Cluster algorithm (MCL) from patient networks or Kmeans from raw data. We then track the identified clusters over ages to construct cluster-trajectories. We compared our novel approaches with three longitudinal clustering approaches by calculating the silhouette score. As a use-case, we analyzed antithrombotic drugs prescribed from 2008 to 2018 contained in the Échantillon Généraliste des Bénéficiaires (EGB), a French national cohort.

**Results:** Our cluster-tracking approaches allowed us to identify several cluster-trajectories having clinical significance. Silhouette score comparison between the different approaches reveals that the best score is obtained for the cluster-tracking approaches.

**Conclusion:** The cluster-tracking approaches are a novel and efficient alternative to identify patient clusters from medico-administrative databases by taking into account their specificities.

## 1 INTRODUCTION

The reuse of medico-administrative databases is nowadays extremely popular. Such databases are indeed increasingly available for epidemiological, clinical and healthcare research to study a large range of health-related issues [1]. However, medico-administrative databases are complex and appropriate analysis methods are required [2]. First, each patient is described through a large number of variables. Analysis methods able to deal with high dimensional data are hence needed. Second, these variables are of a different nature (e.g., drug prescriptions, diagnoses, hospitalizations), and the methods need to consider heterogeneity. Finally, the variables vary over time and are measured over different follow-up periods, thereby generating truncated data when focusing on a given stage of life or disease.. This time dimension is very difficult to apprehend and, overall, only few methods can deal with high dimensional truncated longitudinal data.

Among the various objectives targeted by the reuse of medico-administrative databases, the identification of clusters (i.e., subgroups) of patients is particularly significant. Indeed, given the complexity and the heterogeneity of human diseases, we have to move from a “one size fits all” paradigm towards a more personalized care and a better understanding of disease heterogeneity [3, 4].

To the best of our knowledge, three categories of approaches are available to cluster patients using longitudinal data. These longitudinal clustering approaches are raw-data-based, feature-based and model-based [5]. In raw-data-based approaches, classical (non-longitudinal) clustering algorithms, such as Kmeans, adapt their similarity measure to be applied to the raw longitudinal data. For instance, Kmeans adapted to raw longitudinal data has been used to identify clusters of children based on inattention and hyperactivity during elementary school [6], or to assess the relationships between fibrosis and bioclinical parameters [7]. In feature-based approaches, features are first extracted from the raw longitudinal data. These extracted features are then used as input for classical (non-longitudinal) clustering algorithms. For instance, Wang, Smith, and Hyndman extracted several features from longitudinal data in three (non-clinical) benchmark datasets [8]. They then used the extracted features as input in hierarchical clustering and in an unsupervised neural network algorithm. Although only a small number of features are used for the clustering, the identified clusters are similar to the clusters identified using all the data. Finally, model-based approaches assume that the raw longitudinal data are generated by a mixture of models and intend to extract the parameters of these models. Model-based approaches are, to the best of our knowledge, the most frequently used in biomedical research. The two prevailing model-based approaches are Growth Mixture Modeling (GMM) and Latent Class Growth Analysis (LCGA) [9]. These methods identify clusters of patients based on the common evolution of their longitudinal variables over time. GMM allows small variations around this common evolution between patients within cluster whereas LCGA assumes no variation [10]. Mora et al. applied GMM to identify clusters of women according to the magnitude and timing of depressive symptomatology from pregnancy to two years postpartum [11]. Colder et al. also used GMM to identify clusters of adolescents based on their smoking behavior over four years [12]. LGCA was used by Downie et al. to identify clusters of patients with acute low back pain from pain scores over twelve weeks [13] and by Landa et al. to identify clusters of babies at high risk for autism based on their language, motor and nonverbal cognitive functioning from 6 to 36 months [14].

However, raw-data-based, feature-based and model-based longitudinal clustering approaches have some limitations. For instance, truncated data are not handled. Patients with truncated data must be removed or their data must be imputed. In the context of medico-administrative databases, truncated data are an inescapable issue, as patients are followed-up over a fixed period. In addition, the number of clusters must be specified *a priori*. To determine the optimal number of clusters, criteria are usually used to assess the quality of the clustering [15]. These criteria include for instance the silhouette score [16] [17] or the Davies-Bouldin criterion [18] [19]. However, the optimal number of clusters might differ depending on the criterion chosen [20]. Another limitation specific to the model-based approaches is that the majority of the studies focus on only one longitudinal variable. The joint analysis of two or three longitudinal variables is possible ([21], [22], [23], [24]), but becomes computationally challenging for more than three variables. Finally, in all three categories of approaches, each patient is assigned to only one cluster over the entire time period.

An alternative strategy for clustering patients from longitudinal data could be cluster tracking. Cluster tracking is an approach mainly used in the field of social network analysis [25]. It is a two-step strategy. In the first step, the clusters are identified at each time point. In the second step, the clusters are matched between the different time points to allow their tracking along the timeline. Clusters are identified at each time point using non-longitudinal clustering algorithms [26, 27].

Different methods can be used to identify clusters including widely used methods such as Kmeans or more recent approaches such as network clustering algorithms.. For instance, Li et al. constructed a patient network based on clinical similarity and performed a clustering approach in order to identify subtypes of type 2 diabetes [28]. Wang et al. constructed patient networks from omics data and identified clusters of cancer patients with different survival profiles [29]. Patient networks have the advantage of preserving privacy because the interactions between patients are considered rather than absolute data [30]. In addition, a large number of algorithms exist for clustering networks [31]. However, to our knowledge, current network-based approaches to identify patient clusters do not consider longitudinal data.

We propose here novel cluster-tracking approaches to identify patient clusters and trajectories from longitudinal data contained in medico-administrative databases. Our approaches starts by identifying clusters of patients at each age. We compared the performance of such method using two clustering strategies: Kmeans directly applied to the raw data or the Markov Cluster algorithm (MCL) applied to patient networks constructed from raw data. We then track the clusters identified at the different ages based on their sharing of patients. As a use-case, we analyzed drug prescriptions of the national cohort managed by the French health insurance, called the Échantillon Généraliste des Bénéficiaires (EGB). We identified different trajectories of patient clusters with clinical interest. Finally, we compared these cluster-tracking approaches with three existing types of longitudinal clustering approaches, by calculating a modified silhouette score. The best modified silhouette scores were obtained with the two cluster-tracking approaches.

## 2 MATERIAL AND METHODS

### 2.1 Cluster-tracking approach

We propose novel approaches for clustering patients from longitudinal data extracted from medico-administrative databases. These approaches start by identifying clusters of patients at each considered age. To this goal, we used two different clustering strategies: the Markov Cluster algorithm (MCL) applied to patient networks built from raw data and Kmeans applied directly on raw data. Clusters are then tracked over ages to define cluster-trajectories.

#### 2.1.1 Identifying clusters of patients using MCL from patient networks

The first clustering strategy used to identify clusters of patients relies on the construction of patient networks. We started by constructing a patient network for each age considered. We then applied the MCL clustering algorithm on each network.

##### Constructing patient networks

A patient network is a graph *G* = (*V, E*) with *V* patient nodes and *E* edges representing interactions between patient nodes. We built a network for each patient age. Each network is constructed using a similarity matrix 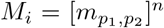 where *n* is the number of patients, *i* is the age and 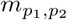 is the similarity between patients *p*_1_ and *p*_2_ having the same age. This similarity matrix is symmetrical, with 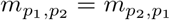.

The similarity between patients at age *i* can be computed using different similarity measures. We tested four different similarity measures: the Cosine similarity, the opposite of the normalized Euclidean distance, the Jaccard index and the generalized Jaccard index (*Supplementary section S1*).

The similarity matrices built for each considered age are then filtered according to a threshold *t*. The goal of the filtering step is to obtain networks with a reduced number of edges [32]. The filtered matrices are next used to build patient networks. We tested different thresholds. For each threshold *t*, the filtered matrix 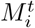 is obtained as follows:

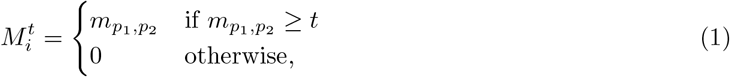

where a null value indicates that patients *p*_1_ and *p*_2_ have a similarity value below the threshold *t* and will thereby not be connected in the patient network. From each similarity matrix 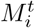, the associated patient network can be constructed. An edge between patients *P*_1_ and *P*_2_ is weighted by the value 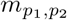 of the matrix.

Reducing the number of edges may lead to disconnected nodes. Therefore, we selected the threshold *t* in the similarity matrices which allowed us to obtain the minimum number of isolated patient nodes in any network (*Supplementary section S2*).

##### Clustering patient networks

We applied the Markov Cluster algorithm (MCL) [33] on the largest connected component of the patient networks. The MCL algorithm uses random walks to simulate flows on the network. The flows allow to distinguish network areas where nodes are strongly connected, which correspond to the clusters. We used the version 0.0.6.dev0 of the “markov-clustering” Python package with the default parameters.

#### 2.1.2 Identifying clusters of patients using Kmeans from raw data

We described in the previous section a clustering strategy based on patient networks using MCL. We also used Kmeans as a second clustering strategy. Kmeans is applied directly on raw data, for each age considered. In Kmeans, the number of clusters must be specified *a priori*. We determined the optimal number of clusters per age by calculating the silhouette score [34]. The silhouette score assesses the clustering quality by computing the separation distance between the obtained clusters.

Let us define

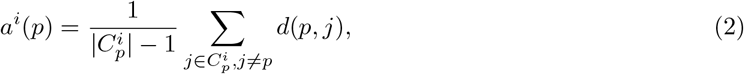

the mean distance of patient *p* to their cluster 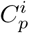 at age *i*, with 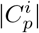 the number of patients in 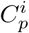 and *d*(*p, j*) the Euclidean distance between patients *p* and *j* belonging to 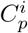, and let

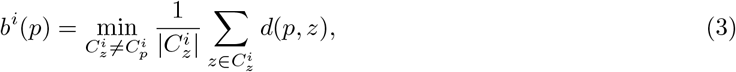

be the mean distance of a patient *p* to their neighboring cluster 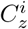 at age *i*, with 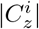 the number of patients in 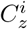 and *d*(*p, z*) the Euclidean distance between the patient *p* belonging to 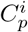 and the patient *z* belonging to 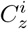.

We start by calculating the silhouette score for each patient of age *i* as follows:

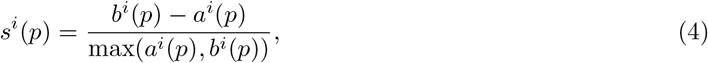

The silhouette score at a given age *i* over all the patients is obtained as follows:

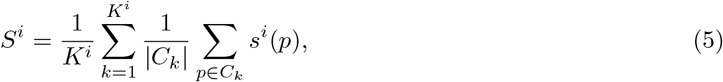

with *K*^*i*^ the number of clusters at age *i*, |*C*_*k*_| the number of patients in the cluster *C*_*k*_.

The silhouette score varies between -1 and 1. Values close to 1 indicate that the clusters are well-separated. Values close to 0 indicate overlapping clusters. Negative values indicate that a patient is assigned to a wrong cluster.

#### 2.1.3 Tracking the clusters over ages

In the previous step, we identified sets of clusters per age either from patient networks with MCL or from raw data with Kmeans. We then intend to follow the clusters over the different ages. Let *C*^*i*^ and *C*^*i*+1^ be two sets of clusters identified at 2 consecutive ages, *i* and *i* + 1. We computed the intersection (i.e., the number of common patients) between every pair of clusters (*c, c*^′^) obtained at 2 consecutive ages:

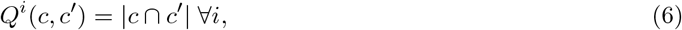

with *c* ∈ *C*^*i*^ and *c*^′^ ∈ *C*^*i*+1^.

Next, for each cluster *c* ∈ *C*^*i*^, we identified the cluster from the set of clusters *C*^*i*+1^ having the greatest number of common patients as follows:

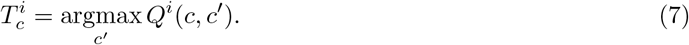

Please note that if, for the cluster *c*, there is more than one cluster match in 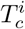 (i.e., if there is more than one cluster with the same maximum number of common patients), all the clusters are included in 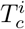.

We visualized the tracking of clusters with an alluvial plot, in which the blocks represent the clusters and the stream fields between the blocks represent the number of common patients. The height of the blocks and the thickness of the stream fields are proportional to the number of patients.

#### 2.1.4 Identifying cluster-trajectories

We identified in the previous section sets of successive clusters. We called the sets of successive clusters cluster-trajectories. Patients in the same cluster-trajectory are considered to follow the same evolution over time for the longitudinal variables of interest.

The cluster-trajectories are visualized using a flowchart composed of blocks representing the clusters. The arrow thickness between the blocks represents the number of common patients. All clusters identified are described using the meta-information available for the patients.

### 2.2 Longitudinal clustering approaches

We compared the performance of the cluster-tracking approaches proposed in this work to existing state-of-the-art approaches dedicated to clustering patients using longitudinal data. The three categories of state-of-the-art longitudinal clustering approaches are raw-data-based, feature-based and model-based approaches [5, 35]. We selected three specific methods, each representative of a category of approach. All longitudinal clusters identified with these methods are described using the meta-information available for the patients.

#### 2.2.1 Raw-data-based approach

Raw-data-based approaches work directly with longitudinal raw data [5, 35]. We selected Kml3d, an R package providing an implementation of Kmeans specifically designed for longitudinal data [36]. This package takes as input a 3-dimensional matrix *M* (*n, i, y*) with *n* the patients, *i* the age and *y* the set of variables characterizing the patients. The algorithm calculates the Euclidean distance between all patients (in n-dimensional space). Patients with the smallest distance are grouped in the same cluster. Importantly, the number of cluster needs to be defined *a priori*.

Kml3d cannot handle truncated data but allows imputation using different methods. We used the copy mean method (default), which imputes truncated data using a linear interpolation and adds a variation to adapt the shape of the interpolation to the shape of the mean of the other values [37]. Patients are removed from the analysis when their number of truncated data are greater than |*I*| − 2, with *I* the set of patient ages.

#### 2.2.2 Feature-based approach

Raw data usually have a high dimension. The goal of the feature-based approaches is to reduce the dimensions by extracting several features characterizing the longitudinal data [5, 35]. These features can then be used as input in classic (non-longitudinal) clustering algorithms, such as Kmeans or hierarchical clustering. We extracted the most common features: mean, standard deviation, kurtosis and skewness [38]. The kurtosis and the skewness describe the shape of the distribution of longitudinal data. We therefore obtained four features per patient and per longitudinal variable. These features were used as input in Kmeans.

#### 2.2.3 Model-based approach

In model-based approaches, each longitudinal variable is characterized by a model or a mixture of models [5, 35]. We applied Growth Mixture Modeling (GMM), which assumes that a model with a given mean and shape is associated with each cluster [10]. Let *y*_*p*_ be a longitudinal variable of the patient *p* composed of *j* repeated observations and *K* the number of clusters, distributed with probabilities *π*_*k*_ with *k* = 1, …, *K, π*_*k*_ ∈ [0, 1] and Σ_*k*_ *π*_*k*_ = 1. A growth mixture modeling is defined as follows:

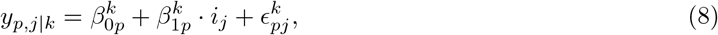

with *i*_*j*_ the patient’s age at the j*th* observation of the variable 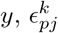 the time-specific residual errors, and 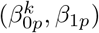 the patient-specific coefficients.

In GMM, analyzing several variables simultaneously is computationally challenging. GMM can be applied separately for each variable, but this assumes that all longitudinal variables are independent from each other.We hence decided to use an aggregated variable 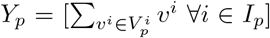, with *I*_*p*_ the set of ages of the patient *p* and 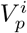 the set of longitudinal variables of the patient *p* at age *i*. This aggregated variable allows us to apply a single GMM.

GMM calculates for every patient their posterior probability of belonging to each cluster using this aggregated variable as input. The cluster assigned to each patient is the one with the greatest posterior probability.

#### 2.2.4 Determining the optimal number of clusters

In the raw-data-based, the feature-based and the model-based approaches, the number of clusters must be specified as a parameter *a priori*. In order to determine the optimal number of clusters, we calculated several classic clustering quality criteria (*Supplementary section S3*). In the raw-data-based and the feature-based approaches, we calculated the Calinski-Harabasz criterion [39], the Kryszczuk variant of Calinski-Harabasz criterion [40], the Genolini variant of Calinski-Harabasz criterion [36], the opposite of Ray-Turi criterion [41] and the opposite of Davies-Bouldin criterion [42]. In the model-based approach, we calculated the Akaike Information Criterion (AIC) [43] and the Bayesian Information Criterion (BIC) [44]. Furthermore, for all the approaches, we calculated a modified silhouette score as follows:

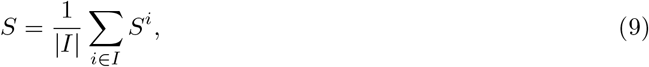

with *S*^*i*^ the silhouette score at the age *i* (equation 5) and *I* the set of patient ages. In this modified silhouette score, we calculated the silhouette score *S*^*i*^ at each age rather than over the entire period. This avoids imputing truncated data.

### 2.3 Choice of the metric to compare the performances of the different approaches

In the cluster-tracking approaches, we used two clustering strategies: one based on network using MCL (section 2.1.1) and one based on raw data using Kmeans (section 2.1.2). In order to compare the clustering quality of these two clustering strategies, we calculated the modified silhouette score (equation 9). We also calculated this modified silhouette score in the three longitudinal-clustering approaches. This allowed us to compare the clustering quality of the different approaches.

We estimated the 95% confidence interval of the modified silhouette score using the percentile bootstrap method [45]. We generated 100 bootstrap samples by resampling with replacement patients present in the population of interest. In each bootstrap sample, we applied the different approaches and we calculated the modified silhouette score. We obtained the confidence interval by taking the 2.5*th* and the 97.5^*th*^ percentile of the distribution of the modified silhouette scores.

### 2.4 Use-case: the Echantillon Généraliste des Bénéficiaires

We used longitudinal health data from the Echantillon Généraliste des Bénéficiaires (EGB), a French medico-administrative database. The EGB is a random sample from the French health insurance database [46]. It is representative of the French population and contains approximately 660,000 individuals followed over a period of 11 years. We confirm that this study has been declared to INSERM (Institut National de la Santé et de la Recherche Médicale, https://www.inserm.fr/). The information provided to individuals in EGB on the possible re-use of their data and the procedures for exercising their rights comply with the legislative and regulatory provisions applicable to the processing of personal data in the SNDS. According to French regulation, individuals in SNDS database are informed of the reuse of their data for research and can opposed to this reuse as defined by Articles 92 to 95 of Decree No. 2005-1309 of 20 October 2005 (https://www.legifrance.gouv.fr/loda/article_lc/LEGIARTI000037300884/).

As required from French regulation, EGB data can be reuse for research projects from authorized persons once the research project is declared to their institution (INSERM).

Among others, EGB contains drug reimbursements, which are longitudinal high dimensional data that can be used to identify subgroups of patients (*Figure 1*). We extracted data on drugs prescribed between 2008 and 2018. For each patient, the date of prescription, the Anatomical Therapeutic Chemical (ATC) class and the name of the prescribed drugs are indicated (see example *Table 1*). The ATC class is an international classification of drugs established by the World Health Organization (WHO) [47]. We only considered prescribed drugs belonging to the ATC class of antithrombotic agents (i.e., B01). We obtained 164,942 patients with such prescriptions. We further selected patients aged 60 to 70 and having had at least one drug prescription for two or more consecutive months. Our goal was to focus only on patients with sustained prescriptions. Our final dataset is composed of 30,111 different patients and 19 different drugs. There is a majority of men in this population, with a sex ratio (men/women) of 0.61. This is consistent with the fact that cardiovascular diseases, which accounts for the majority of antithrombotic prescriptions, is more common in men.

**Figure 1:**
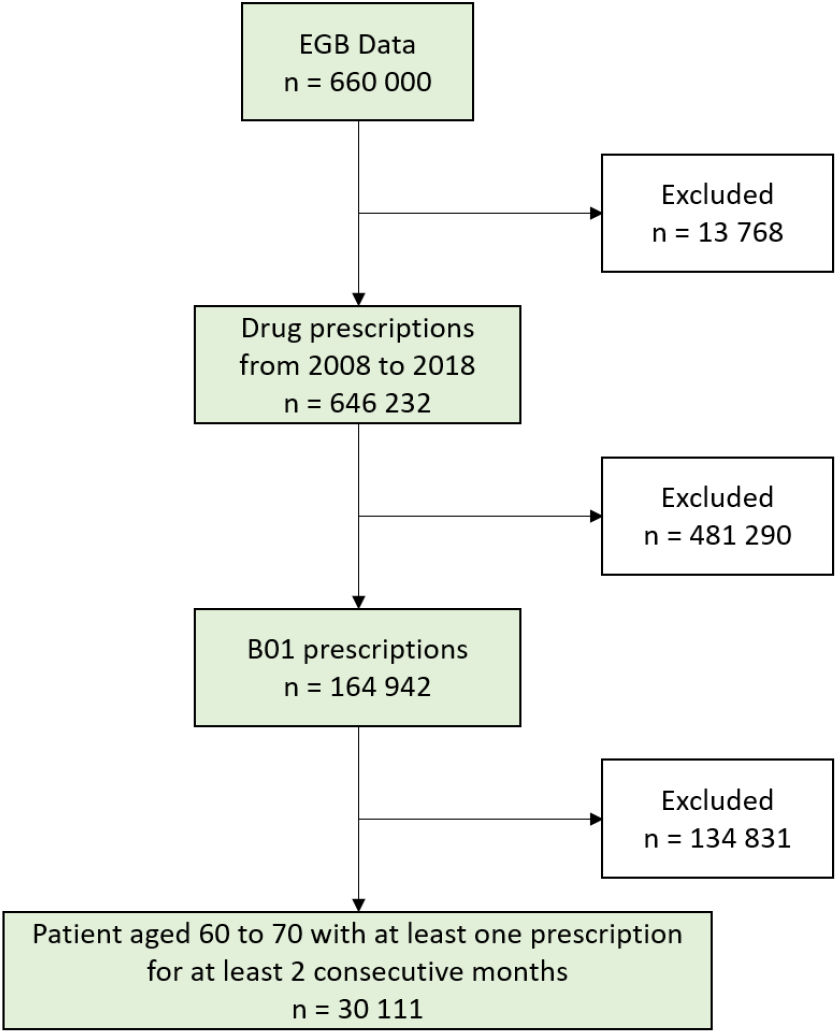
Extraction of longitudinal data from the EGB, considered as a use-case in this study From the EGB medico-administrative database, we extracted antithrombotic drugs prescribed for at least two consecutive months from 2008 to 2018 in patients ages 60 to 70. n: number of patients, B01: antithrombotic agents

**Table 1:** Example of drug prescriptions contained in the EGB M01: Anti-inflammatory and antirheumatic products, B01: antithrombotic agents, N02: Analgesics

| Patient ID | Prescription date | ATC class | Drug name |
| --- | --- | --- | --- |
| $P_1$ | 01/04/2008 | M01 | Ibuprofen |
| $P_1$ | 01/12/2015 | B01 | Aspirin |
| $P_2$ | 01/02/2010 | N02 | Tramadol |
| $P_3$ | 01/05/2016 | B01 | Clopidogrel |

We also extracted data on long-term illnesses (i.e., illnesses that last at least 6 months) from the EGB. 23,063 patients out of the 30,111 patients studied experienced at least 1 long-term illness between 60 and 70 years old. These long-term illnesses represent 865 distinct diseases. Each disease is coded with the 10*th* revision of the international statistical classification of diseases and related health problems (ICD-10 code).

We calculated, for each patient, the sum of the prescriptions per drug at a given age (see example *Table 2*). We therefore obtained a table per patient age. Focusing on patients aged 60 to 70 years old, we obtained a total of 11 tables.

**Table 2:** Example of total number of prescriptions per drug calculated for three drugs and three patients aged 60 years

| Patient ID | B01_1 | B01_2 | B01_3 |
| --- | --- | --- | --- |
| $P_1$ | 0 | 10 | 5 |
| $P_2$ | 1 | 8 | 4 |
| $P_3$ | 2 | 6 | 3 |

Importantly, we observed three types of truncated data (*Figure 2*).

**Figure 2:**
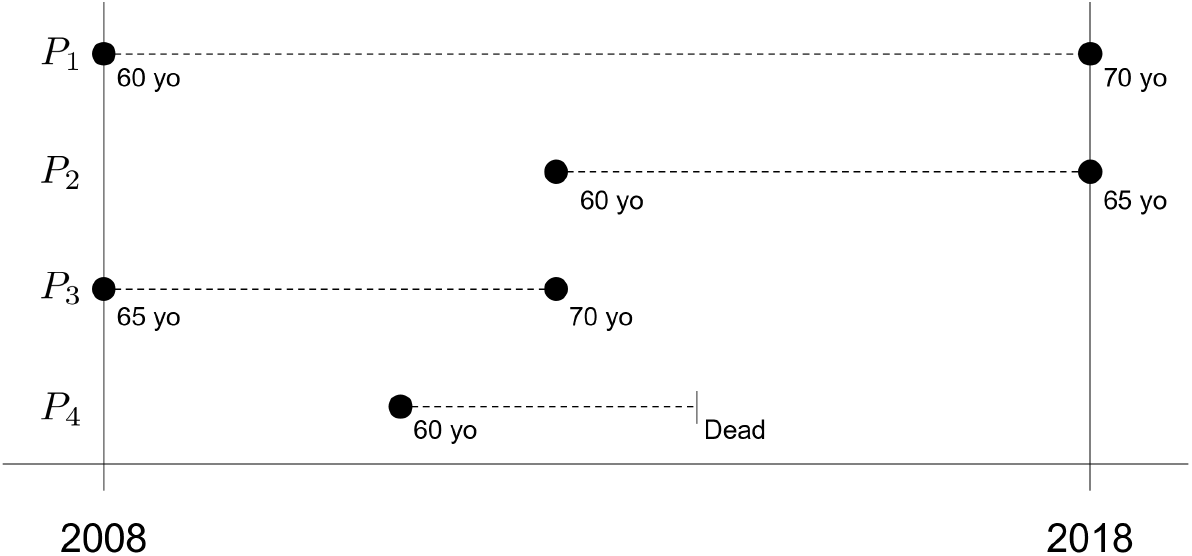
Example of patient follow-up in the EGB *P*_1_ has no truncated data. Truncated data is observed when a patient is 60 years old after 2008 (Patient *P*_2_), when a patient is 60 years old before 2008 (Patient *P*_3_) and when a patient dies before the end of the follow-up (Patient *P*_4_). A patient can be associated with several types of truncated data. For example, the patient *P*_4_ is 60 years old after 2008 and dies before the end of the follow-up.

## 3 RESULTS

### 3.1 Cluster-tracking approaches allow identifying and tracking patient clusters over ages to identify cluster-trajectories

We first apply two different clustering strategies to identify clusters of patients at each age considered in our use case. The first clustering strategy is applied to patient networks (Material and methods 2.1.1). The second clustering strategy is directly applied to raw data (Material and methods 2.1.2). The clusters are then tracked over ages to define cluster-trajectories.

#### 3.1.1 Identifying cluster-trajectories with a cluster-tracking approach based on networks using MCL

The first clustering strategy used in the cluster-tracking approach relies on the construction of patient networks (Material and methods 2.1.1). Patient networks are constructed using similarity matrices. Different measures can be computed to calculate similarities between patients and construct the similarity matrices (*Supplementary section S1*). We selected the Cosine similarity because it has the greatest variance. Using this Cosine similarity, we constructed 11 similarity matrices. In each matrix, the similarities are computed between all patients of a given age (from 60 to 70 years old). Patient networks are then constructed by applying a threshold on the similarity matrices. Patients associated with a similarity higher than the threshold will be linked by an edge in the patient network. We tested different Cosine similarity thresholds and selected a threshold of 0.8. This threshold was chosen as the best trade-off to minimize number of isolated patients while reducing the number of edges (*Supplementary section S2*). We obtained 11 patient networks (one by age, see *Table 3* and *Figure 3* for the network of patients aged 60 years old).

**Table 3:**
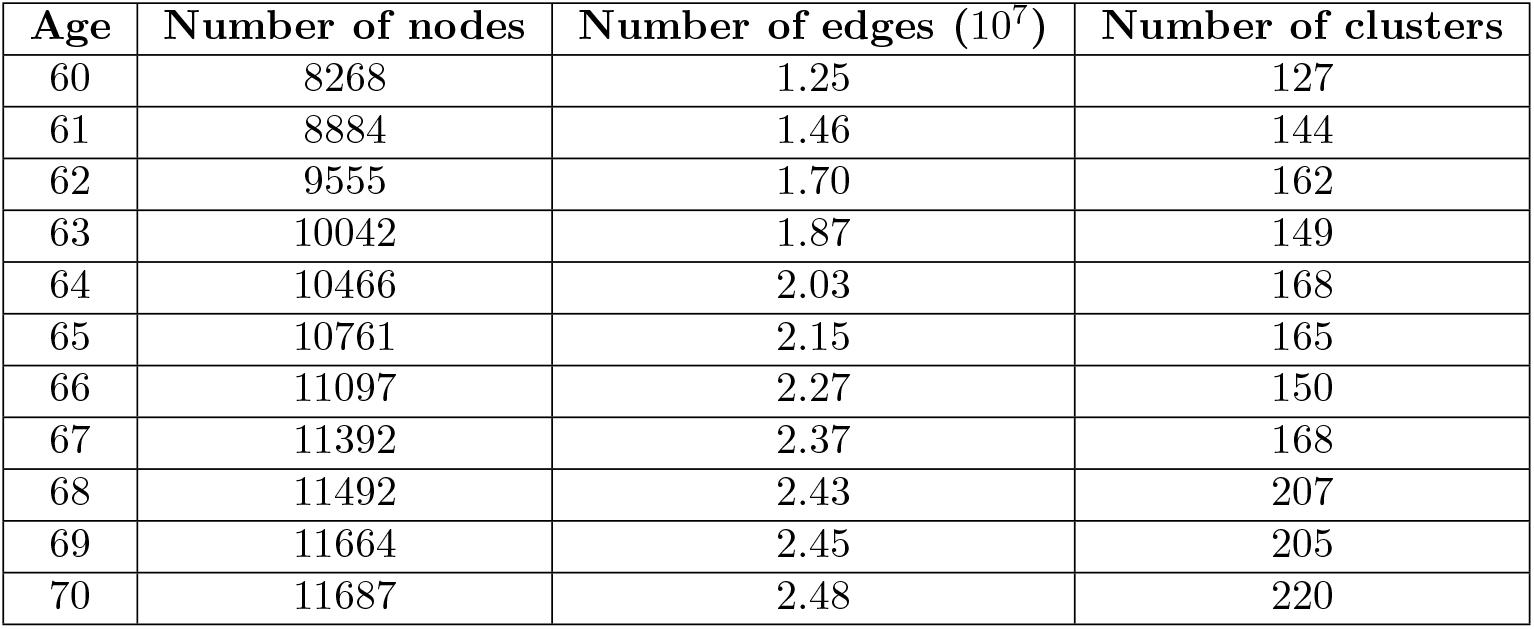
Number of nodes, edges and clusters in 60 to 70 years old patient networks

**Figure 3:**
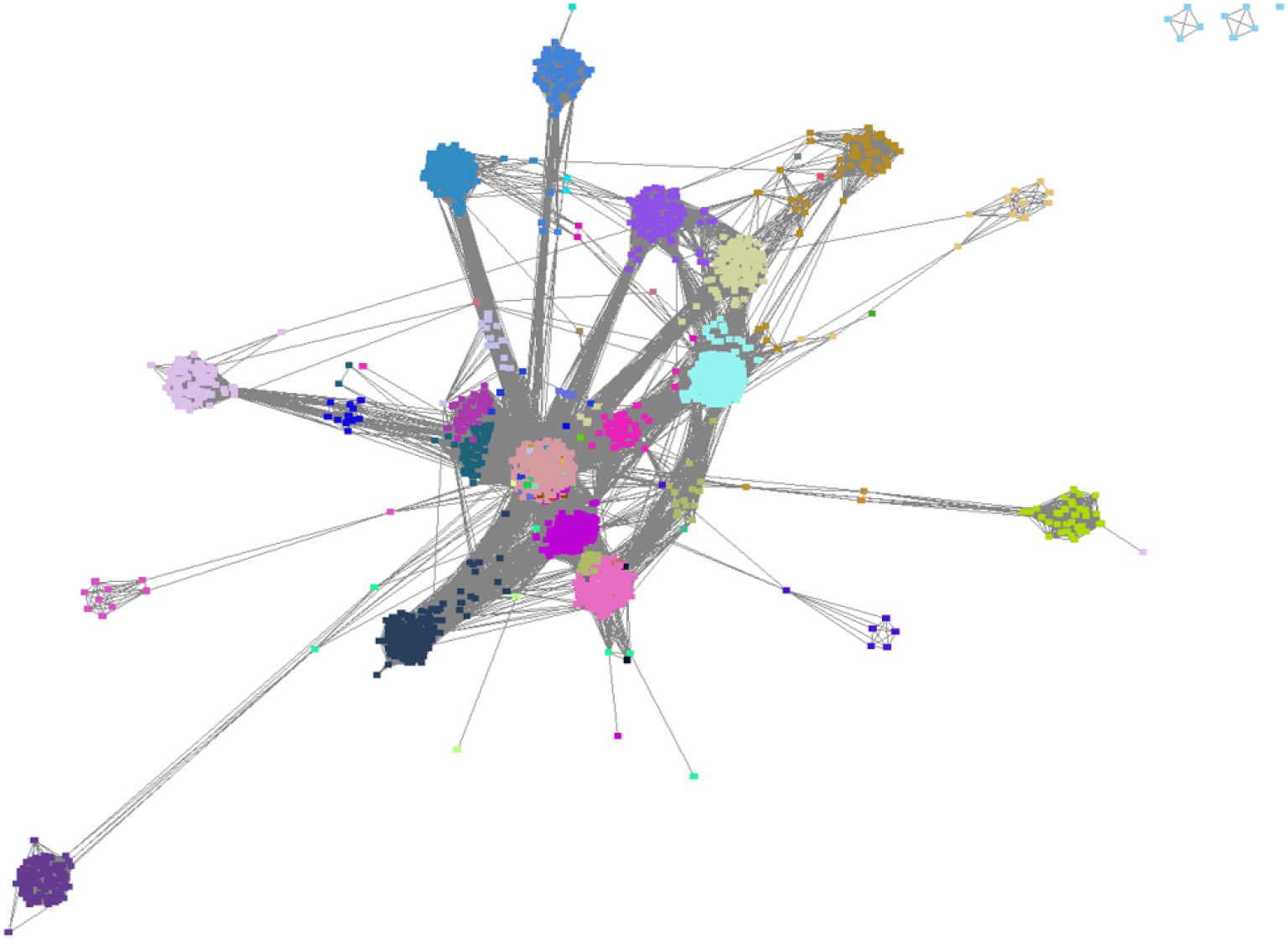
60-year-old patients network Nodes represent patients and edges the interactions between patients having a Cosine similarity of at least 0.8. The length of edges is inversely proportional to the Cosine similarity. Nodes of the same color belong to one of the 127 clusters identified with the Markov Cluster algorithm.

We then applied the Markov Cluster algorithm (MCL) to identify clusters of patients (Material and methods 2.1.1). The MCL algorithm is applied systematically on all the 11 patient networks, revealing different numbers of clusters per network (*Table 3*). For example, in the patient network constructed at 60 years old, 127 clusters are identified (*Figure 3*).

We next computed the number of common patients between clusters identified at consecutive ages (Material and methods 2.1.3). This allows tracking the evolution of the clusters over consecutive ages (*Figure 4*) and identifying cluster-trajectories. We identified 12 cluster-trajectories composed of clusters with at least 100 patients (*Supplementary section S4*). We described the clusters that compose these trajectories with the number of patients, the sex ratio, the two most prescribed drugs and the two most frequent long-term illnesses. Most of the 12 identified trajectories are composed of clusters with a majority of men. This is explained by the presence of a majority of men in our study population (i.e., 30,111 patients). Indeed, the sex ratio of this population is 0.61.

**Figure 4:**
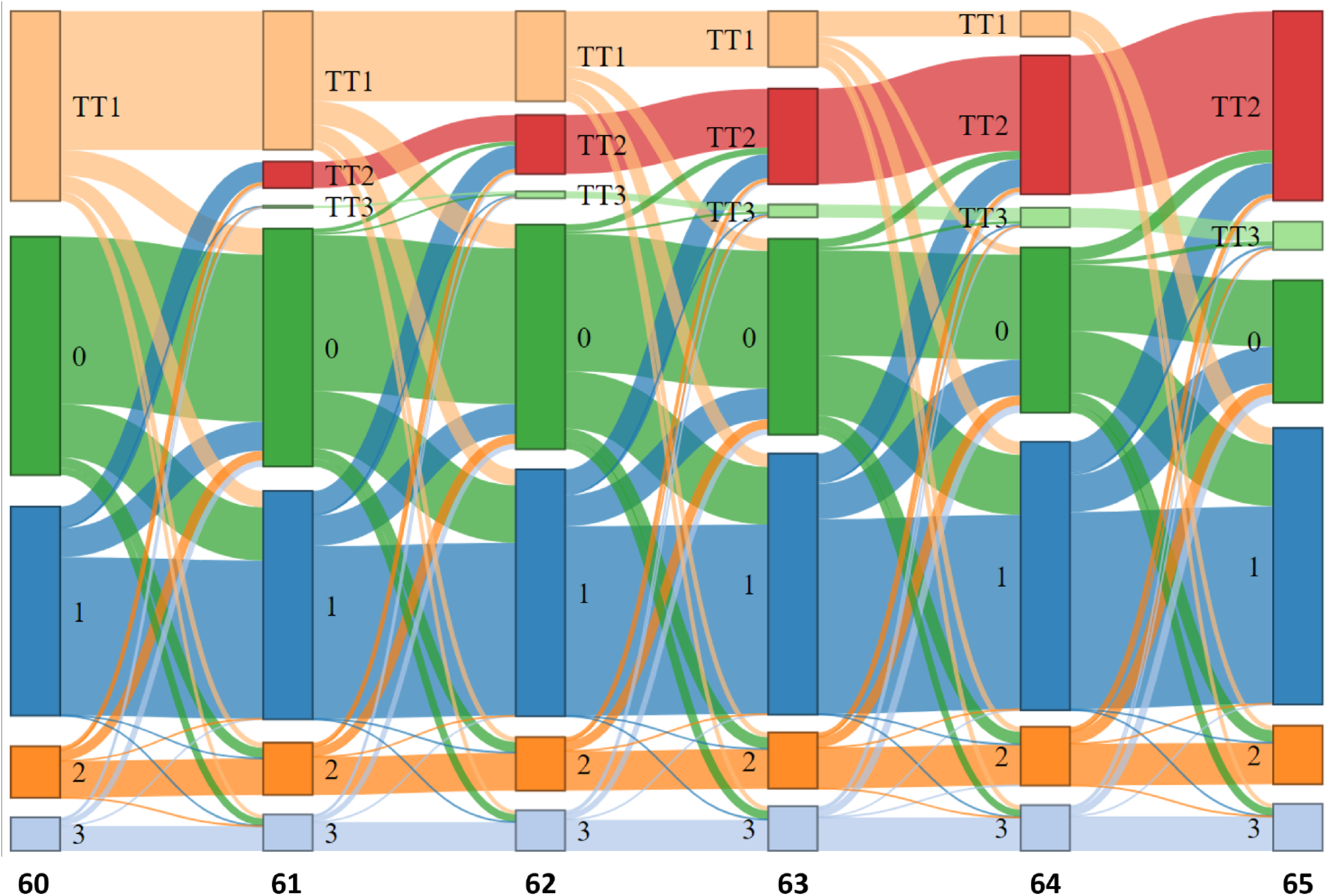
Tracking of clusters identified from patient networks In the alluvial plot, the blocks represent the clusters obtained with the Markov Cluster algorithm from networks constructed at each patient age. The stream fields between blocks represent the number of common patients. The height of the blocks and the thickness of the stream fields are proportional to the number of patients. For the sake of clarity, we have only represented the cluster tracking from 60 to 65 years old. We focus on the three largest clusters identified with the MCL algorithm (clusters 1 to 3, numbered from the largest to the smallest). Cluster 0 represents patients with no prescription at the given age. The three types of truncated data are represented in clusters TT1 (Truncated Type 1), TT2 (Truncated Type 2) and TT3 (Truncated Type 3). Clusters TT1 contains patients aged 60 after 2008; Clusters TT2 contains patients aged 60 before 2008 and Clusters TT3 contains patients who have died before the end of follow-up (*Figure 2*).

We next focused on the 3 cluster-trajectories (A,B and C) with the largest number of patients (*Figure 5* and *Supplementary section S4*). The trajectory A is the one with the largest number of patients. By analyzing clusters of this trajectory, we observed that aspirin is prescribed to all patients at all ages. Furthermore, more than half of the patients present in any cluster of the trajectory A are also present in the following cluster. For instance, among the 4238 patients of the cluster identified at age 60, 3 209 (i.e., 76 %) are present in the cluster of age 61. Thus, for the majority of the patients, aspirin is prescribed for at least two consecutive years. In addition, at 63 and 64 years old, two clusters are observed in the trajectory A. The first cluster is associated with aspirin prescription only and the second cluster is associated with enoxaparin prescription in addition to aspirin. These two clusters merge into the same cluster at the following age, and aspirin is the only drug prescribed in the merged cluster. This implies that, when enoxaparin is prescribed in addition to aspirin, the majority of the patients switch to aspirin-only prescriptions the following year. The most frequent long-term illnesses observed in clusters that compose this trajectory is diabetes (ICD-10 code E11). This diagnosis is also observed in all the 12 trajectories identified. No specific anti-thrombotic drugs are recommended for patients suffering from diabetes. However, diabetes increases cardiovascular risk and therefore many patients with antithrombotic drugs have diabetes [48]. The other long-term illness observed in the trajectory A is chronic ischemic heart disease (ICD-10 code I25). Indeed, antithrombotic therapy is a key part of secondary prevention in patients with chronic ischemic heart disease and patients with this illness are considered for long-term aspirin treatment [49].

**Figure 5:**
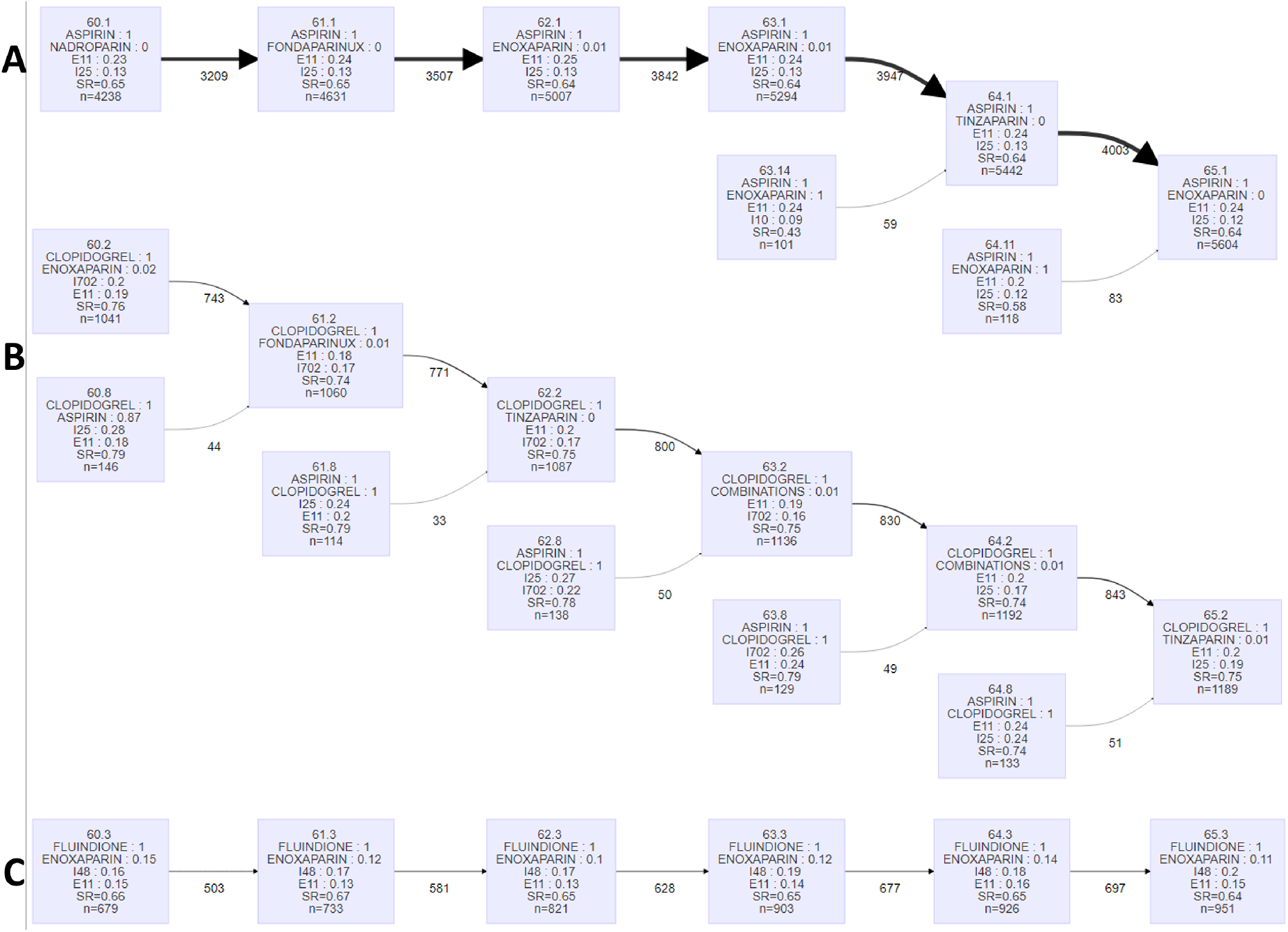
Subset of patient cluster-trajectories identified with the cluster-tracking approach based on network We represented 3 cluster-trajectories (A,B and C) out of the 12 identified. We represented them from 60 to 65 years old. In these 3 cluster-trajectories, each box represents a cluster. Each cluster is named as follows: “x.y”, with x the age at which it was identified and y its cluster number in the alluvial plot (*Figure 4*). The clusters are characterized by the two most prescribed drugs (name, percentage of patients receiving the drug), the two most frequent long-term illnesses (ICD-10 code, percentage of patients), the sex ratio (SR) and the total number of patients (n). The number under arrows is the number of common patients between the two blocks. The arrow thickness is proportional to this number. Combinations: combinations of two platelet aggregation inhibitors. ICD-10 code E11: type 2 diabetes mellitus, I25: chronic ischemic heart disease, I10: essential primary hypertension, I702: atherosclerosis of arteries of extremities, I48: atrial fibrillation.

The trajectory B is composed of clusters in which clopidogrel, an antiplatelet drug, is prescribed to all patients at all ages. Two clusters are systematically observed at each age. In the first cluster, clopidogrel is the only drug prescribed. In the second cluster, aspirin is prescribed in addition to clopidogrel. These two clusters merge into the same cluster at the following age. In the merged cluster, clopidogrel is the only drug prescribed. Hence, we can observe that when aspirin is prescribed in addition to clopidogrel, the majority of the patients switch to clopidogrel-only the following year. After myocardial infarction (ICD-10 code I25), that we observed in clusters with clopidogrel-only prescription, and percutaneous coronary intervention, dual antiplatelet is recommended during several months followed by a switch to mono-therapy [50]. In the trajectory B, the most frequent long-term illness in addition to diabetes is peripheral arterial disease (ICD-10 code I702). Antiplatelet therapy is indicated in patients with peripheral arterial disease that are symptomatic or have undergone revascularization. Clopidogrel is the preferred antiplatelet drug in this case [51].

The trajectory C is composed of clusters of patients with fluindione prescriptions; about 12 % of the patients also have enoxaparin prescriptions. More than half of the patients present in any cluster of the trajectory C are also present in a cluster of the following year. For instance, among the 679 patients present in the cluster identified at age 60, 503 (i.e., 74 %) are present in the cluster identified at the age 61. Thus, we can conclude that, for the majority of the patients, fluindione is prescribed for at least two consecutive years. The most frequent long-term illness in this trajectory is atrial fibrillation (ICD-10 code I48). Fluindione, which is a vitamin K antagonist, has been shown to strongly reduce stroke in patients with atrial fibrillation [52]. In the recent period, non-vitamin K antagonist oral anticoagulants have been recommended in replacement of vitamin K antagonists such as apixaban and rivaroxaban [53]. These two drugs have the same indication. Because non-vitamin K antagonist oral anticoagulants are more convenient to use, the switch of drugs observed from age 67 is consistent (*Supplementary section S4*).

The same interpretations were carried out for the 9 remaining cluster-trajectories (*Supplementary section S4*). In each cluster that compose these trajectories, we always observe a drug prescribed to all patients (i.e., predominant drug). Most of the time, more than half of the patients present in the clusters of these trajectories are also present in the following-age clusters. Thus, the predominant drugs are usually prescribed for at least two consecutive years. However, this is not the case in the cluster-trajectory D. In this trajectory, two types of clusters are usually observed at each age. The first cluster contains enoxaparin prescriptions to all the patients and the second cluster contains tinzaparin prescriptions to all the patients. These two clusters systematically merge into the cluster 0 at the following age. The cluster 0 is composed of patients with no antithrombotic prescriptions. Thus, the majority of patients with enoxaparin or tinzaparin prescriptions in this trajectory no longer have prescriptions at the following year. Enoxaparin and tinzaparin are low molecular weight heparin. Hence, we hypothesize that we captured patients having an acute venous thromboembolism event in this trajectory. This cluster-trajectory D is also the only one with clusters composed of a majority of women (i.e., sex ratio about 0.40). Associated comorbidities are scarce, with the most frequent diagnoses being cancers (ICD-10 codes C50, C34, C18), for which there is a known significant increase of thromboembolism event requiring low molecular weight heparin [54]. Moreover it is well-known that women have a higher risk of thromboembolism event than men [55].

#### 3.1.2 Identifying cluster-trajectories with the cluster-tracking approach based on raw data using Kmeans

In the previous section (3.1.1), we identified cluster-trajectories using a network-based cluster-tracking approach. We also implemented a cluster-tracking approach using Kmeans applied to raw data (Material and methods 2.1.2). In this second strategy, we applied a Kmeans per patient age, from 60 to 70 years old.

In Kmeans, the number of clusters must be specified *a priori*. We calculated the silhouette score and identified an optimal number of clusters at each patient age (*Supplementary section S5*). The optimal number of clusters was between 6 and 8. We then tracked the clusters identified by Kmeans over ages (Material and methods 2.1.3). We identified 9 cluster-trajectories composed of clusters with at least 100 patients (*Supplementary section S6*). We described these trajectories with the number of patients, the sex ratio, the two most prescribed drugs and the two most frequent long-term illnesses. We observed that all trajectories are composed of a majority of men. This is explained by the presence of a majority of men in our study population (i.e., 30,111 patients).

For the sake of simplicity, we next focused on three cluster-trajectories (A,B and C). We represented them from 60 to 65 years old (*Figure 6*). The trajectory A is the one with the largest number of patients. Aspirin is prescribed to all patients in the clusters that compose this trajectory. In all the clusters of the trajectory B, clopidogrel is prescribed to all patients. In all the clusters of the trajectory C, fluindione is prescribed to all patients and enoxaparin is prescribed to about 12 % of patients. In addition, more than half of the patients present in any cluster of these three trajectories are also present in the following-age clusters. Thus, we can conclude that, for the majority of the patients, aspirin, clopidogrel and fluindione are prescribed for at least two consecutive years in the trajectories A, B, and C, respectively. As in the network-based cluster-tracking approach, diabetes (ICD-10 code E11) is one of the most frequent long-term illnesses observed in clusters of all identified trajectories. The other long-term illness observed in the trajectory A is chronic ischemic heart disease (ICD-10 code I25). As mentioned previously, long-term aspirin treatment is prescribed to patients with previous myocardial infarction [49]. In trajectory B, the most frequent long-term illness in addition to diabetes in the clusters of age 60 and 61 is peripheral arterial disease (ICD-10 code I702). Clopidogrel is the preferred antiplatelet drug indicated in patients with peripheral arterial disease [51]. In the clusters identified from 62 years old, the most frequent long-term illness is chronic ischemic heart disease (ICD-10 code I25) for which clopidogrel is also recommended and which is associated to peripheral arterial disease because both are arteriopathies. In trajectory C, the most frequent long-term illness is atrial fibrillation (ICD-10 code I48). As mentioned previously, fluindione has been shown to strongly reduce stroke in patients with atrial fibrillation [52].

**Figure 6:**
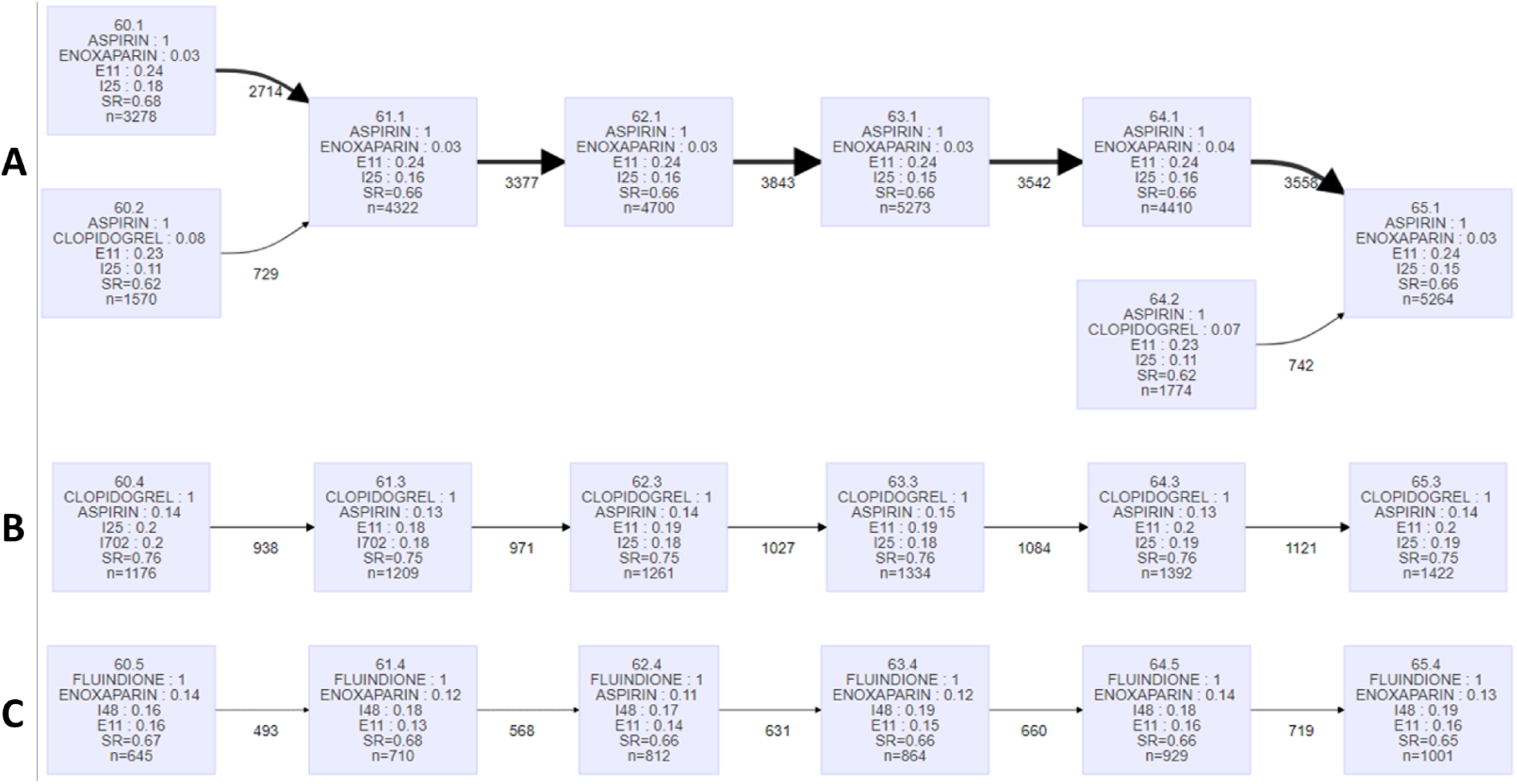
Subset of patient cluster-trajectories identified with the raw-data-based cluster-tracking approach We represented 3 cluster-trajectories out (A,B and C) of the 9 identified. We represented them from 60 to 65 years old. In these 3 trajectories, each block represents a cluster. Each cluster is named as follows: “x.y”, with x the age time at which it was identified and y the number of the cluster. The clusters are characterized by the two most prescribed drugs (name, percentage of patients), the two most frequent long-term illnesses (ICD-10 code, percentage of patients), the sex ratio (SR) and the number of patients (n). The number under arrows is the number of patients in common between the two blocks. The arrow thickness is proportional to this number. ICD-10 code E11: type 2 diabetes mellitus, I25: chronic ischemic heart disease, I702: atherosclerosis of arteries of extremities, I48: atrial fibrillation.

These same descriptions were carried out for the 6 other cluster-trajectories (*Supplementary section S6*). In each cluster that compose these trajectories, we always observe a predominant drug prescribed to all patients. Hence, we can conclude that the predominant drug is prescribed for at least two consecutive years. This is not the case in the cluster-trajectories D and F. In the trajectory D, several clusters merge into the cluster 0, which is composed of patients with no antithrombotic prescriptions. Thus, most of the patients in this trajectory no longer have prescriptions at the following year. Contrarily to what we previously observed in the network-based cluster-tracking approach, this trajectory D is not composed of a majority of women (i.e., sex ratio about 0.40). In the trajectory F, combinations (i.e., combinations of two platelet aggregation inhibitors) are prescribed to all patients in clusters identified from 61 to 67 years old. Then aspirin is prescribed to about 60% of patients in clusters identified from 68 years old. As hemorrhage risk increases with age, patients at older age switch to only one platelet aggregation inhibitor.

### 3.2 Comparing the two clustering strategies used in the cluster-tracking approaches

We identified the cluster-trajectories with the cluster-tracking approaches using two different clustering strategies: one based on the construction of patient networks by applying the MCL algorithm and one based on raw data by applying Kmeans. We aimed to compare the performances of these two clustering strategies.

We observed that the trajectories A in the two cluster-tracking approaches are composed of clusters having a similar description (*Supplementary sections S4 and S6*). Indeed, aspirin is prescribed to all the patients and the two most frequent long-term illness are the sames in all the clusters. We also observed a similar description between the clusters of the trajectories C and E of the two cluster-tracking approaches. The clusters of the two trajectories G also have a similar description, but the two trajectories do not begin at the same age. The first cluster is identified at 60 years old with the network-based cluster-tracking approach and at 64 years old with the raw-data-based cluster-tracking approach. The two trajectories H also begin at different ages. In both cases, the cluster-trajectories identified with the network-based cluster-tracking approach start at earlier ages than the cluster-trajectories identified with the raw-data-based cluster-tracking approach.

We calculated the modified silhouette score (*S*) and its 95% confidence interval to assess clustering quality in the two cluster-tracking approaches (Material and methods 2.3). We obtained *S* = 0.50 ([0.46 ; 0.55]) with the network-based cluster-tracking approach and *S* = 0.57 ([0.53 ; 0.58]) with the raw-data-based cluster-tracking approach (*Table 4 B*.). *A priori*, the cluster-tracking approach seems to be more efficient using a raw-data-based than a network-based strategy. However, the confidence intervals overlap.

**Table 4:** Longitudinal-clusters identified with the three longitudinal clustering approaches and comparison with the cluster-tracking approaches A. n: number of patients, SR: sex ratio (percentage of men), Top 2 drugs: the two most prescribed drugs with the percentage of patients, Top 2 diseases: the two most frequent long-term illnesses (ICD-10 code) with the percentage of patients. In all approaches, the identified longitudinal-clusters are ranked from the largest to the smallest. Combinations: combinations of two platelet aggregation inhibitors. ICD-10 codes C34: malignant neoplasm of bronchus and lung, C50: malignant neoplasm of breast, C61: malignant neoplasm of prostate, E11: type 2 diabetes mellitus, I10: essential primary hypertension, I21: acute myocardial infarction, I25: chronic ischemic heart disease, I702: atherosclerosis of arteries of extremities, I48: atrial fibrillation, K74: fibrosis and cirrhosis of liver. B. silhouette scores calculated in the different approaches and their 95% confidence intervals.

|  | Raw-data-based longitudinal-clustering |  |  |  | Feature-based longitudinal-clustering |  |  |  | Model-based longitudinal-clustering |  |  |  |
| --- | --- | --- | --- | --- | --- | --- | --- | --- | --- | --- | --- | --- |
|  | n | SR | Top 2 drugs (%) | Top 2 diseases (%) | n | SR | Top 2 drugs (%) | Top 2 diseases (%) | n | SR | Top 2 drugs (%) | Top 2 diseases (%) |
| A | 12550 | 0.53 | Aspirin (65)<br>Enoxaparin (22) | E11 (23)<br>I25 (7) | 11510 | 0.62 | Aspirin (100)<br>Enoxaparin (11) | E11 (32)<br>I25 (17) | 14461 | 0.65 | Aspirin (76)<br>Clopidogrel (27) | E11 (28)<br>I25 (18) |
| B | 8665 | 0.64 | Aspirin (100)<br>Enoxaparin (15) | E11 (32)<br>I25 (21) | 7484 | 0.51 | Aspirin (41)<br>Enoxaparin (28) | E11 (17)<br>I48 (6) | 6822 | 0.50 | Aspirin (52)<br>Enoxaparin (23) | E11 (19)<br>I10 (6) |
| C | 2937 | 0.75 | Clopidogrel (100)<br>Aspirin (60) | E11 (29)<br>I25 (28) | 2827 | 0.73 | Clopidogrel (100)<br>Aspirin (44) | E11 (27)<br>I25 (24) | 2481 | 0.77 | Aspirin (92)<br>Clopidogrel (66) | I25 (44)<br>E11 (29) |
| D | 1794 | 0.65 | Fluindione (99)<br>Enoxaparin (43) | I48 (24)<br>E11 (22) | 2460 | 0.62 | Aspirin (100)<br>Enoxaparin (20) | E11 (31)<br>I25 (15) | 2198 | 0.59 | Aspirin (74)<br>Enoxaparin (18) | E11 (29)<br>I10 (10) |
| E | 1013 | 0.65 | Rivaroxaban (70)<br>Aspirin (41) | I48 (37)<br>E11 (23) | 2050 | 0.60 | Fluindione (100)<br>Enoxaparin (38) | I48 (24)<br>E11 (19) | 2033 | 0.55 | Aspirin (70)<br>Fluindione (17) | E11 (25)<br>I10 (8) |
| F | 402 | 0.81 | Combinations (100)<br>Aspirin (79) | I25 (45)<br>E11 (28) | 1114 | 0.74 | Clopidogrel (100)<br>Aspirin (89) | I25 (41)<br>E11 (30) | 1296 | 0.61 | Aspirin (82)<br>Enoxaparin (22) | E11 (30)<br>I25 (13) |
| G | 345 | 0.77 | Ticagrelor (98)<br>Aspirin (97) | I25 (46)<br>I21 (33) | 615 | 0.77 | Aspirin (100)<br>Clopidogrel (100) | I25 (37)<br>E11 (31) | 803 | 0.58 | Aspirin (78)<br>Fluindione (18) | E11 (31)<br>I10 (12) |
| H | 243 | 0.62 | Warfarin (100)<br>Enoxaparin (46) | E11 (23)<br>I48 (19) | 576 | 0.62 | Rivaroxaban (100)<br>Aspirin (16) | I48 (32)<br>E11 (16) | 15 | 0.80 | Aspirin (100)<br>Clopidogrel (87) | I25 (67)<br>E11 (13) |
| I | 233 | 0.64 | Acenocoumarol (99)<br>Enoxaparin (42) | E11 (23)<br>I48 (18) | 509 | 0.80 | Combinations (100)<br>Aspirin (97) | I25 (51)<br>E11 (33) | 2 | 1.00 | Aspirin (100)<br>Fondaparinux (50) | C61 (50)<br>K74 (50) |
| J | 106 | 0.82 | Prasugrel (96)<br>Aspirin (93) | I25 (62)<br>E11 (28) | 454 | 0.70 | Fluindione (100)<br>Aspirin (96) | E11 (28)<br>I48 (22) |  |  |  |  |
| K | 73 | 0.66 | Apixaban (82)<br>Aspirin (33) | I48 (30)<br>E11 (18) | 410 | 0.63 | Rivaroxaban (100)<br>Aspirin (60) | I48 (34)<br>E11 (23) |  |  |  |  |
| L | 13 | 0.85 | Ticlopidine (100)<br>Aspirin (54) | E11 (23)<br>C34 (15) | 102 | 0.62 | Warfarin (100)<br>Aspirin (61) | E11 (26)<br>I25 (17) |  |  |  |  |

Table 4: Longitudinal-clusters identified with the three longitudinal clustering approaches and comparison with the cluster-tracking approaches
| Network-based cluster-tracking | Raw-data-based cluster-tracking | Raw-data-based longitudinal-clustering | Feature-based longitudinal-clustering | Model-based longitudinal-clustering |
| --- | --- | --- | --- | --- |
| 0.50<br>[0.46 ; 0.55] | 0.57<br>[0.53 ; 0.58] | 0.27 | 0.20<br>[-0.02 ; 0.20] | -0.33<br>[-0.26 ; 0.01] |

### 3.3 Comparing the cluster-tracking approach with the longitudinal-clustering approaches

We compared the performance of the cluster-tracking approaches based on network and raw-data with three methods representative of the three types of longitudinal clustering approaches, namely raw-data-based, feature-based and model-based approaches (Material and methods 2.2). We used the same longitudinal data extracted from EGB in patients aged from 60 to 70 years old in all the approaches.

#### 3.3.1 Choosing the optimal number of clusters

In the three longitudinal-clustering approaches, the number of clusters need to be specified *a priori*. In order to select an optimal number of clusters, we calculated several classic clustering quality criteria (Material and methods 2.2.4). These criteria however do not point to clear optimums (*Supplementary section S3*). Hence, we next tried to use the modified silhouette score. We also failed to find a clear optimum with this approach. Indeed, the greatest silhouette scores (i.e., global maximum) was obtained for the smallest number of clusters (*Supplementary section S3*). We therefore decided to specify the number of clusters as 12 clusters. This number corresponds to the number of cluster-trajectories identified with the network-based cluster-tracking approach.

#### 3.3.2 Identifying clusters with the raw-data-based longitudinal clustering approach

We applied Kml3d [36], the selected raw-data-based longitudinal clustering approach (Material and methods 2.2.1) to the longitudinal data extracted from the EGB. First, 1737 patients are removed by the Kml3d algorithm because they have more than 9 truncated data (which is the limit with 11 different ages). We applied the Kml3d algorithm with 12 clusters as parameter and we described all the identified longitudinal-clusters with the number of patients, the sex ratio, the two most prescribed drugs and the two most frequent long-term illnesses.

Among the 12 longitudinal-clusters identified by Kml3d, 10 are composed of at least 100 patients (*Table 4 A*.). At least one of the two most prescribed drugs is prescribed to more than 60 % of patients. For instance, aspirin, clopidogrel, combinations, warfarin and ticlopidine are prescribed to all patients in longitudinal-clusters B, C, F, H and L, respectively. Each longitudinal-cluster identified is therefore characterized by a drug that is predominantly prescribed to patients. More than 20 % of patients have diabetes (ICD-10 code E11) in all the longitudinal-clusters except in the longitudinal-cluster G. Atrial fibrillation (ICD-10 code I48) is one of the two most frequent long-term illnesses in longitudinal-clusters D, E, H, I and K. In these clusters, at least 70 % of patients have vitamin K antagonist prescriptions (such as fluindione, warfarin or acenocoumarol) or non-vitamin K antagonist oral anticoagulants prescriptions (such as rivaroxaban or apixaban). As mentioned in the cluster-tracking approach, vitamin K antagonist has been shown to strongly reduce stroke in patients with atrial fibrillation [52]. In the recent period, non-vitamin K antagonist oral anticoagulants have been recommended in replacement to vitamin K antagonists [53]. Chronic ischemic heart disease (ICD-10 code I25) is always observed in the longitudinal-clusters when aspirin is one of the two most prescribed drugs. As mentioned previously, long-term aspirin treatment is prescribed to patients with previous myocardial infarction [49].

Our goal is to compare the 12 longitudinal-clusters obtained with the raw-data-based longitudinal clustering approach with the cluster-trajectories identified with the cluster-tracking approaches. At least one of the two most prescribed drugs is prescribed to more than 60 % of patients in all the clusters that compose the cluster-trajectories (*Supplementary sections S4* and S6) and in all the longitudinal-clusters (*Table 4 A*.). This is not the case in the raw-data-based-cluster-trajectory D where aspirin is prescribed to about 38 % of patients and enoxaparin is prescribed to about 16 % of patients. Therefore, the majority of cluster-trajectories and longitudinal-clusters are characterized by a predominantly prescribed drug. These trajectories and longitudinal-clusters are composed of a majority of men except in the network-based-cluster-trajectory D where the sex ratio is about 0.40. Breast cancer (ICD-10 code C50) is usually one of the two most frequent long-term illness in the clusters that compose the network-based-cluster-trajectory D. Several cluster-trajectories and longitudinal-clusters have a common drug description. For instance, aspirin and enoxaparin are both prescribed in the longitudinal-cluster B and in the cluster-trajectories A of the two cluster-tracking approaches. The two most frequent long-term illnesses are also the same. Conversely, the raw-data-based longitudinal clustering approach is the only one to have identified three longitudinal-clusters characterized by prescriptions of ticagrelor-aspirin, prasugrel-aspirin and ticlopidine-aspirin (G, J and L respectively in *Table 4 A*.). Similarly, the network-based cluster-tracking approach is the only one to have identified cluster-trajectories characterized by prescriptions of enoxaparin-tinzaparin, aspirin-fluindione and dabigatran-enoxaparin (D, J and L respectively in *Supplementary section S4*). Therefore, additional information are given with the raw-data-based longitudinal clustering approach and the network-based cluster-tracking approach compared to the raw-data-based cluster-tracking approach. Furthermore, we calculated the modified silhouette score (*S*) in the raw-data-based longitudinal clustering approach and in the cluster-tracking approaches to compare the clustering quality (Material and methods 2.3). We obtained *S* = 0.27 for the raw-data-based longitudinal-clustering approach, *S* = 0.50 for the network-based cluster-tracking approach and *S* = 0.57 for the raw-data-based cluster-tracking approach (*Table 4 B*.). The 95% confidence intervals of the two strategies of cluster-tracking approach overlap (*Table 4 B*.). Overall, we obtained a better clustering quality with the cluster-tracking approaches compared to the raw-data-based longitudinal clustering approach.

#### 3.3.3 Identifying clusters with the feature-based longitudinal-clustering approach

We extracted 4 standard features from the the antithrombotic drug prescriptions contained in the EGB: the mean, the standard deviation, the kurtosis and the skewness (Material and methods 2.2.2). We therefore obtained a total of 76 features per patient (i.e., 4 features extracted over the 19 antithrombotic drugs). We then used these features as input in Kmeans. Here, the Kmeans clustering is applied over all the ages jointly. As for the raw-data-based longitudinal clustering approach, we applied the Kmeans clustering selecting 12 clusters as parameter. We described the identified longitudinal-clusters with the number of patients, the sex ratio, the two most prescribed drugs and the two most frequent long-term illnesses.

The 12 longitudinal-clusters identified with the feature-based longitudinal clustering approach are all composed of at least 100 patients (*Table 4 A*.). One of the two most prescribed drugs is always prescribed to all patients except in the cluster B. In this cluster, aspirin is prescribed to 41 % of the patients and enoxaparin is prescribed to 28 % of the patients. The majority of the identified longitudinal-clusters is therefore characterized by a predominantly prescribed drug. At least 15 % of patients have diabetes (ICD-10 code E11) in all the clusters. Chronic ischemic heart disease (ICD-10 code I25) is always observed in the clusters where aspirin is one of the two most prescribed drugs. As mentioned previously, long-term aspirin treatment is prescribed to patients with previous myocardial infarction [49].

We compared the 12 longitudinal-clusters obtained in the feature-based longitudinal clustering approach with the cluster-trajectories identified in the cluster-tracking approaches (*Supplementary sections S4* and S6). We observe that the longitudinal-clusters A and D have a common drug and long-term illness description (*Table 4 A*.). Indeed, aspirin and enoxaparin are both prescribed to a similar proportion of patients and the two most frequent long-term illness are the same (i.e., ICD-10 codes E11 and I25). This type of redundant information is not observed in the cluster-trajectories identified with the two cluster-tracking approaches.

We then calculated the modified silhouette score (*S*) in the feature-based longitudinal-clustering approach to compare the clustering quality with the other clustering approaches (Material and methods 2.3). We obtained *S* = 0.20 for the feature-based longitudinal clustering approach (*Table 4 B*.). This score indicates that patients are less well assigned in clusters with the feature-based longitudinal clustering approach than with the cluster-tracking approach and with the raw-data-based longitudinal clustering approach. The clustering quality is therefore better with the cluster-tracking approaches.

#### 3.3.4 Identifying clusters with the model-based longitudinal-clustering approach

The model-based approach that we applied to the antithrombotic drug prescriptions is GMM (Material and methods 2.2.3). We used an aggregated variable with this algorithm because the simultaneous analysis of several variables is computationally challenging [56]. This aggregated variable is calculated, for each patient, as the total number of drugs prescribed at a given age. As before, we applied GMM selecting 12 clusters as parameter. We described the identified longitudinal-clusters with the number of patients, the sex ratio, the two most prescribed drugs and the two most frequent long-term illnesses.

The GMM algorithm assigns patients to the cluster for which they have the greatest posterior probability of belonging. Although we chose 12 clusters as parameter, none of the patients had a greatest posterior probability of belonging to three out of the 12 selected clusters. Therefore, only 9 model-based-wide-clusters were identified.

The longitudinal-clusters A to G are composed of more than 100 patients (*Table 4 A*.). The two remaining clusters are composed of less than 20 patients. In the 9 longitudinal-clusters, we observed that aspirin is prescribed to more than 50 % of patients. All these longitudinal-clusters are therefore characterized by the same predominantly prescribed drug. Diabetes (ICD-10 code E11) is always one of the two most frequent long-term illness except in longitudinal-cluster I. The longitudinal-cluster I is very small with only two patients. One of the patients has prostate cancer (ICD-10 code C61) and the other has fibrosis and cirrhosis of liver (ICD-10 code K74).

We compared the 9 longitudinal-clusters with the cluster-trajectories identified with the cluster-tracking approaches (*Supplementary sections S4* and S6). The longitudinal-clusters are highly different compared to the cluster-trajectories. Indeed, aspirin is prescribed to a majority of patients in all these longitudinal-clusters, which is not the case in the cluster-trajectories. Furthermore, the diversity of the two most prescribed drugs is lower in the longitudinal-clusters since only aspirin, clopidogrel, enoxaparin, fluindione or fondaparinux are observed. In the cluster-trajectories, other drugs such as warfarin, combinations or rivaroxaban are additionally observed. The model-based longitudinal clustering approach therefore identified longitudinal-clusters where patients are more heterogeneous compared to the cluster-tracking approach.

We then calculated the modified silhouette score (*S*) in the model-based longitudinal clustering approach to compare the clustering quality with the other clustering approaches (Material and methods 2.3). We obtained *S* = −0.33 for the model-based longitudinal clustering approach (*Table 4 B*.). This negative score indicates that patients are assigned to the wrong clusters. The model-based longitudinal clustering approach therefore fails to identify patient clusters. Among all the analyzed approaches, the best clustering quality is obtained with the cluster-tracking approaches.

## 4 DISCUSSION

We proposed here novel approaches based on cluster-tracking for clustering patients from longitudinal data extracted from medico-administrative databases. We applied our approaches to the analysis of antithrombotic drug prescriptions extracted from the EGB between 2008 and 2018 in patients aged from 60 to 70 years old. We showed that cluster-tracking approaches are efficient to identify patient trajectories from medico-administrative databases while taking into account the longitudinal, multidimensional and truncated nature of data.

We compared these new cluster-tracking approaches with three classical longitudinal clustering approaches. Using a modified silhouette score, we showed that the cluster-tracking approaches have a higher performance than these classical approaches. This higher performance of the cluster-tracking approaches might arise from the fact that information available for the analysis is increased. Indeed, for all the longitudinal clustering approaches, it was necessary to impute truncated data or remove patients associated with too much truncated data. This is a critical limitation as, in medico-administrative databases, patients are followed at different moments of their life or disease and the number of time points available for each patient may be very different. The cluster-tracking approaches allow avoiding these imputations. In addition, in the cluster-tracking approaches, no patient are removed from the analysis.

Another interesting feature of the cluster-tracking approaches is that patients can switch clusters as their age progresses. A patient can therefore belong to several cluster-trajectories. This allows considering some uncertainty in patient clustering compared to the longitudinal-clustering approaches where a patient belong to a single longitudinal-cluster.

Among cluster-tracking approaches, the strategies based on network and raw data obtained similar performance. However, the network-based cluster-tracking approach did not require to pre-specify the number of clusters, which might be a parameter difficult to set-up.

Importantly, the network-based cluster-tracking approach have also the advantage of preserving privacy because the interactions between patients are considered rather than absolute data. Another advantage is the flexibility of this approach as many different measures can be used to compute the similarity between patients. These similarity measures can then be tuned depending on the data and question at hand. In addition, a large number of algorithms exist for clustering networks.

Overall, the cluster-tracking approaches are a novel and efficient alternative to identify patient clusters from medico-administrative databases by taking into account their specificities. We were able to clinically interpret the identified cluster-trajectories, using their sex ratio, drug prescription and long-term illness data.

## Data Availability

All data simulated are available online at https://github.com/JudithLamb/Cluster-tracking

## 5 CODE AVAILABILITY

The code for our two cluster-tracking approaches is available on GitHub https://github.com/JudithLamb/Cluster-tracking. For privacy reasons, antithrombotic drug prescriptions extracted from the EGB cannot be shared publicly. We hence generated a simulated dataset of 5594 patients with their drug prescriptions from these extracted data. The results obtained from this simulated sample dataset can be visualized in an R Shiny app also available from the GitHub repository.

