## Supplementary Materials for "Tracking patient clusters over time enables to extract all the information available in the medico-administrative databases"

### Tracking clusters of patients built from longitudinal data

### S1 Similarity measures used for constructing patient networks per age

We used a similarity matrix per patient age to construct each patient network. The similarity between patients is calculated with a similarity measure. In this study, we tested four similarity measures: the Cosine similarity, the opposite of the normalized Euclidean distance, the Jaccard index and the generalized Jaccard index. In the following, we consider two sample sets $A$ and $B$ of length $X$ representing the sum of the prescriptions per drug that two different patients had at a given age.

#### S1.1 Cosine similarity

The Cosine similarity between two sample sets $A$ and $B$ calculates the cosine of the angle ($\theta$) between them. It is most commonly used in information retrieval or text mining [1]. This measure is defined as follows:

$$\cos_\theta(A,B) = \frac{A.B}{||A||.||B||} \tag{1}$$

The values range from -1 to 1 , with -1 when the samples are opposite, 0 when samples are different (i.e., orthogonal) and 1 when they are identical.

#### S1.2 Normalized Euclidean distance

In a n-dimensional space, the normalized Euclidean distance between two sample sets $A$ and $B$ is defined as follows [2]:

$$\begin{aligned} NED(A,B) &= \frac{1}{2}.\frac{||(A-\mathbb{E}[A])-(B-\mathbb{E}[B])||^2}{||(A-\mathbb{E}[A])||^2+||(B-\mathbb{E}[B])||^2} \\ &= \frac{1}{2}.\frac{Var(A-B)}{Var(A)+Var(B)}, \end{aligned} \tag{2}$$

with $\mathbb{E}[A]$ and $\mathbb{E}[B]$, the expectation of $A$ and $B$.
By calculating the opposite of the normalized Euclidean distance, values range from 0 when the samples are different to 1 when they are identical.

#### S1.3 Jaccard index

The Jaccard index calculates the similarity between two sample sets $A$ and $B$ by the ratio of their intersection over their union [3]:

$$J(A,B) = \frac{|A\cap B|}{|A\cup B|}, \tag{3}$$

*These authors contributed equally to this work.

When $A$ and $B$ are numeric, we transform each of their element $x_j$ by $x_j^{new} = \mathbb{1}_{x_j \geq 1}$. We then obtain two new sample sets $A' \in \{0, 1\}$ and $B' \in \{0, 1\}$. The Jaccard index between them is defined as follows:

$$J(A', B') = \frac{M_{11}}{X - M_{00}}, \tag{4}$$

with $X$ the length of the two sample sets, $M_{11} = \sum_{i=1}^{X}(\mathbb{1}_{A'_i=1} \times \mathbb{1}_{B'_i=1})$ and $M_{00} = \sum_{i=1}^{X}(\mathbb{1}_{A'_i=0} \times \mathbb{1}_{B'_i=0})$. This index gives a value between 0 when the two samples are different and 1 when they are identical.

#### S1.4 Generalized Jaccard index

The generalized version of the Jaccard index allows to calculates the similarity between two numeric vectors, without transforming their elements. The generalized Jaccard index between two sample sets $A$ and $B$ is defined as follows [4]:

$$Jg(A, B) = \frac{\sum_{i=1}^{X} \min(A_i, B_i)}{\sum_{i=1}^{X} \max(A_i, B_i)} \tag{5}$$

with $X$ the length of sample sets $A$ and $B$. The obtained values are ranges from 0 when the samples are different to 1 when they are identical.

We calculated the Cosine similarity, the Jaccard index, the opposite of the normalized Euclidean distance and the generalized Jaccard index between all patients in our use-case, for each age from 60 to 70 years old. We selected the Cosine similarity for constructing networks because this is the similarity measure having the greatest variance (*Figure S1*). This similarity measure is the one that best distinguishes similar patients from dissimilar patients.

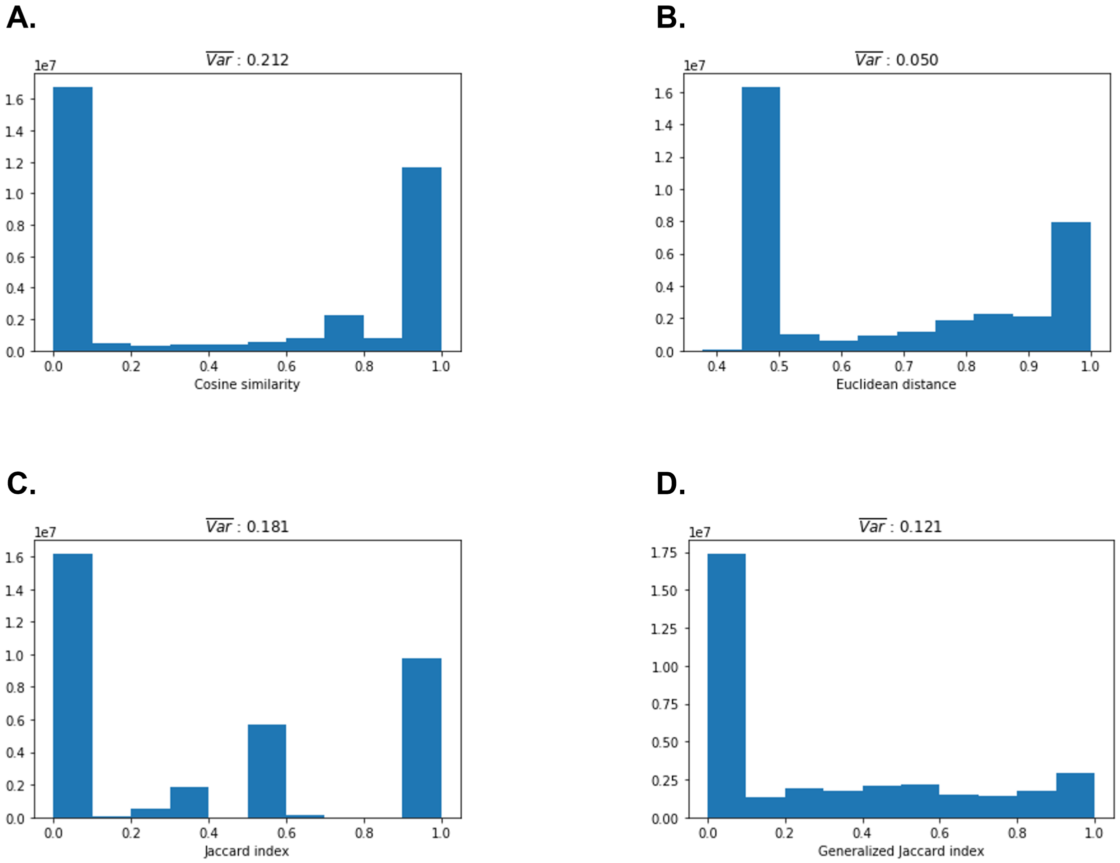


Figure S1: Similarity measure distributions and the mean variance

A: Distribution of the Cosine similarity, B: Distribution of the opposite of the normalized Euclidean distance, C: Distribution of the Jaccard index, D: Distribution of the generalized Jaccard index. The different distributions relate patients aged 60.

$\overline{Var}$: Mean variance from 60 to 70 years old. For each similarity measure, we calculated the distribution variance at each age and we took the mean.

#### S2 Choice of the threshold applied in similarity matrix

We varied the Cosine similarity from 0 to 1 with a step of 0.1 to choose the threshold. For each threshold tested, we calculated the number of edges and the number of isolated patients (i.e., patient connected to none of the other patients) obtained in the associated network (*Figure S2*). The number of isolated patients is under 1% from 0 to 0.9. We selected 0.8 as threshold because this is where we observe the fastest decrease in the number of edges.

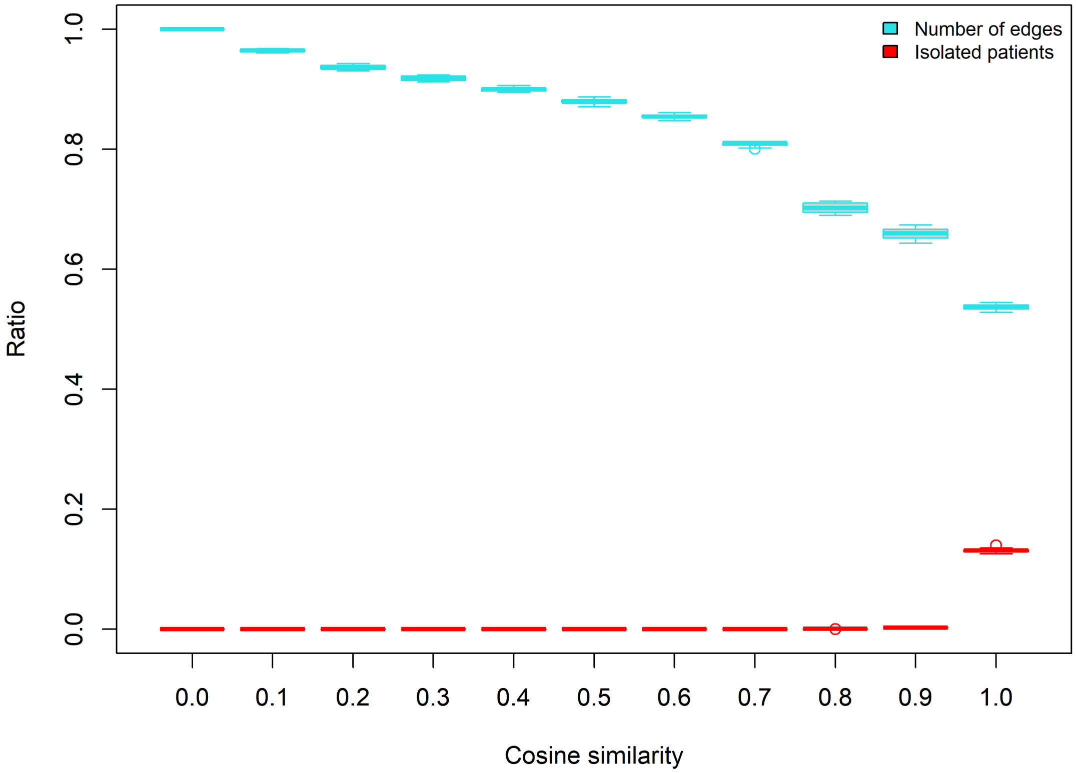


Figure S2: Choice of the Cosine similarity threshold
For each value of Cosine similarity, the blue box represents the number of edges in each similarity matrices from 60 to 70 years old, and the red box represents the isolated patients (i.e., patient connected to none of the other patients)

#### S3 Assessing the optimal number of clusters using several quality criteria in the three longitudinal clustering approaches

The number of clusters must be specified in the three longitudinal clustering approaches. We used several quality criteria to find the optimal number of clusters. Kml3d, the selected raw-data-based longitudinal-clustering approach, computes five different quality criteria to help selecting the optimal number of clusters: Calinski-Harabasz criterion [5], Kryszczuk variant of Calinski-Harabasz criterion [6], Genolini variant of Calinski-Harabasz criterion [7], the opposite of Ray-Turi criterion [8] and the opposite of Davies-Bouldin criterion [9] (*Figure S3 A*). We also used these five quality criteria in the the feature-based longitudinal-clustering approach to find the optimal number of clusters (*Figure S3 B*). We used the Akaike Information Criterion (AIC) [10] and the Bayesian Information Criterion (BIC) [11] in GMM, the selected model-based longitudinal-clustering approach, as they are calculated by the algorithm (*Figure S3 C*). We also calculated in all the longitudinal approaches, the modified silhouette score (*Figure S4*).

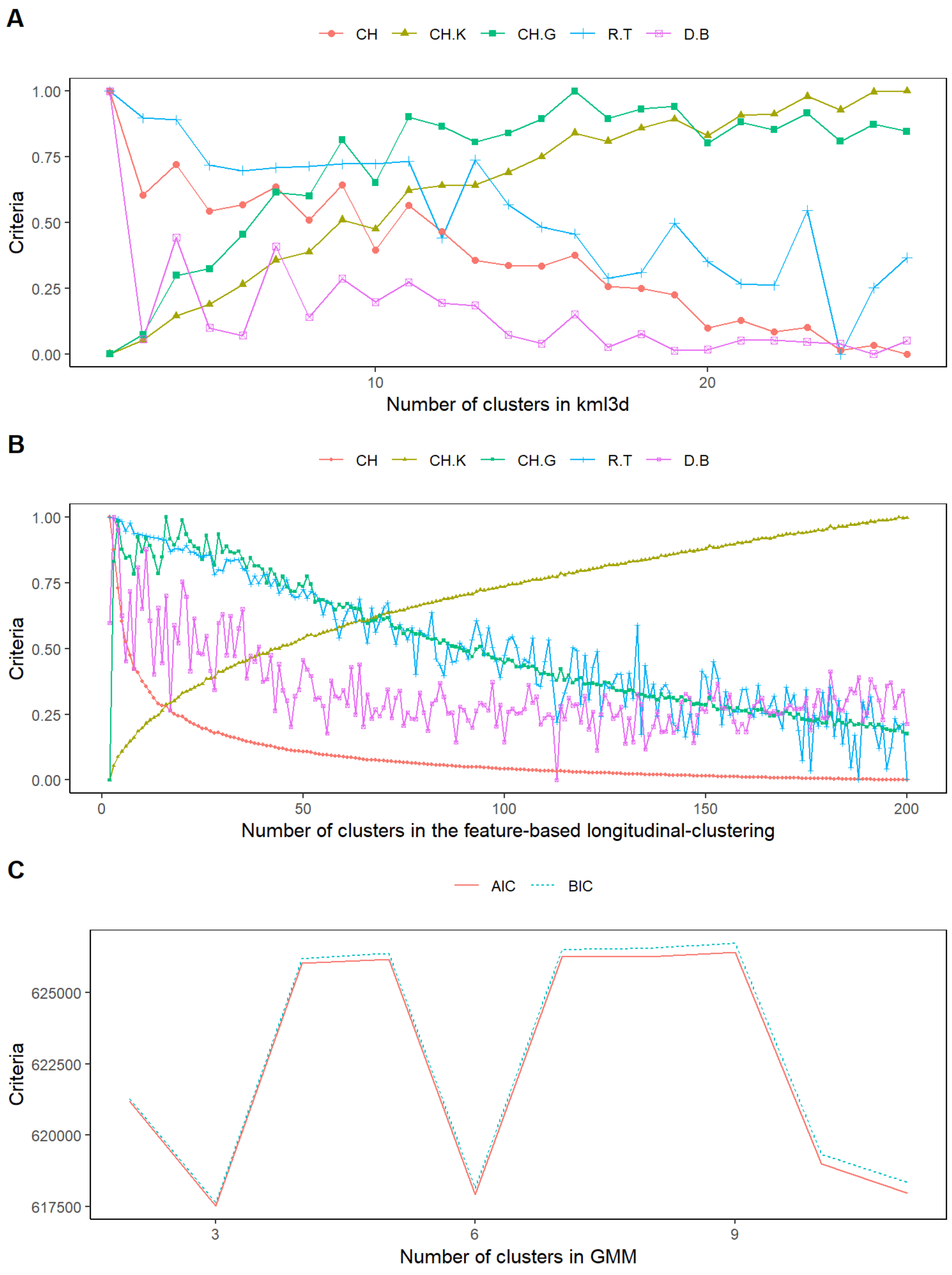


Figure S3: Choice of the optimal number of clusters with several quality criteria computed by the different longitudinal clustering approaches

A: Kml3d, the selected raw-data-based longitudinal-clustering approach, allowed to vary the number of clusters from 2 to 26, B: We varied the number of clusters from 2 to 200 in the feature-based longitudinal-clustering approach, C: We varied the number of clusters only from 2 to 10 in GMM, the selected model-based longitudinal-clustering approach, because of the complexity in the computational time.

CH: Calinski-Harabasz criterion, CH.K: Calinski-Harabasz Kryszczuk variant criterion, CH.G: Calinski-Harabasz Genolini variant criterion, R&T: Ray-Turi criterion, D&B: Davies-Bouldin criterion, AIC: Akaike Information Criterion, BIC: Bayesian Information Criterion

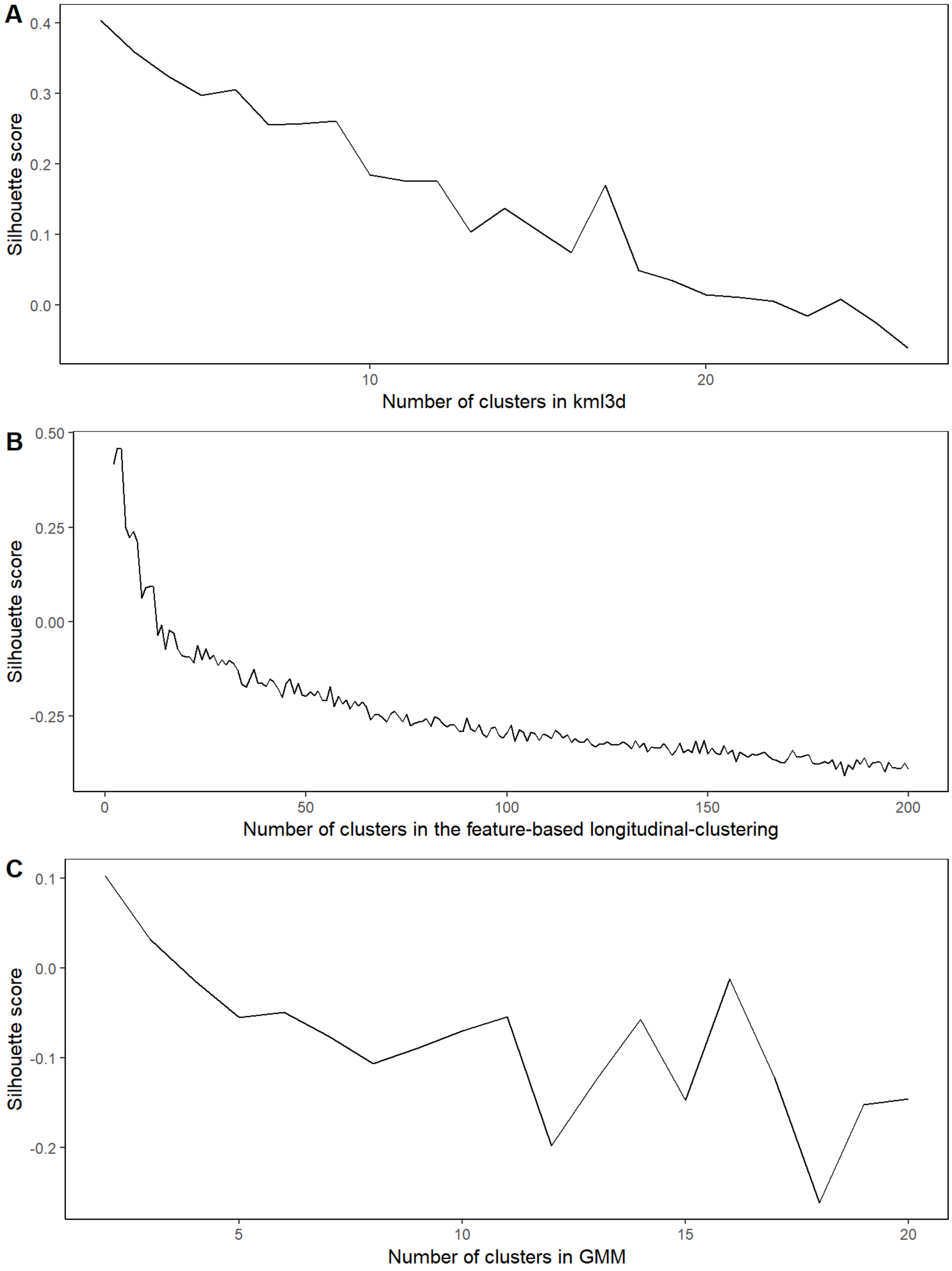


Figure S4: Choice of the optimal number of clusters with the modified silhouette score in the three longitudinal clustering approaches

A: Kml3d, the selected raw-data-based longitudinal-clustering approach, allowed to vary the number of clusters from 2 to 26, B: We varied the number of clusters from 2 to 200 in the feature-based longitudinal-clustering approach, C: We varied the number of clusters only from 2 to 20 in GMM, the selected model-based longitudinal-clustering approach because of the complexity in the computational time (11 days).

#### S4 Cluster-trajectories identified with the network-based cluster-tracking approach

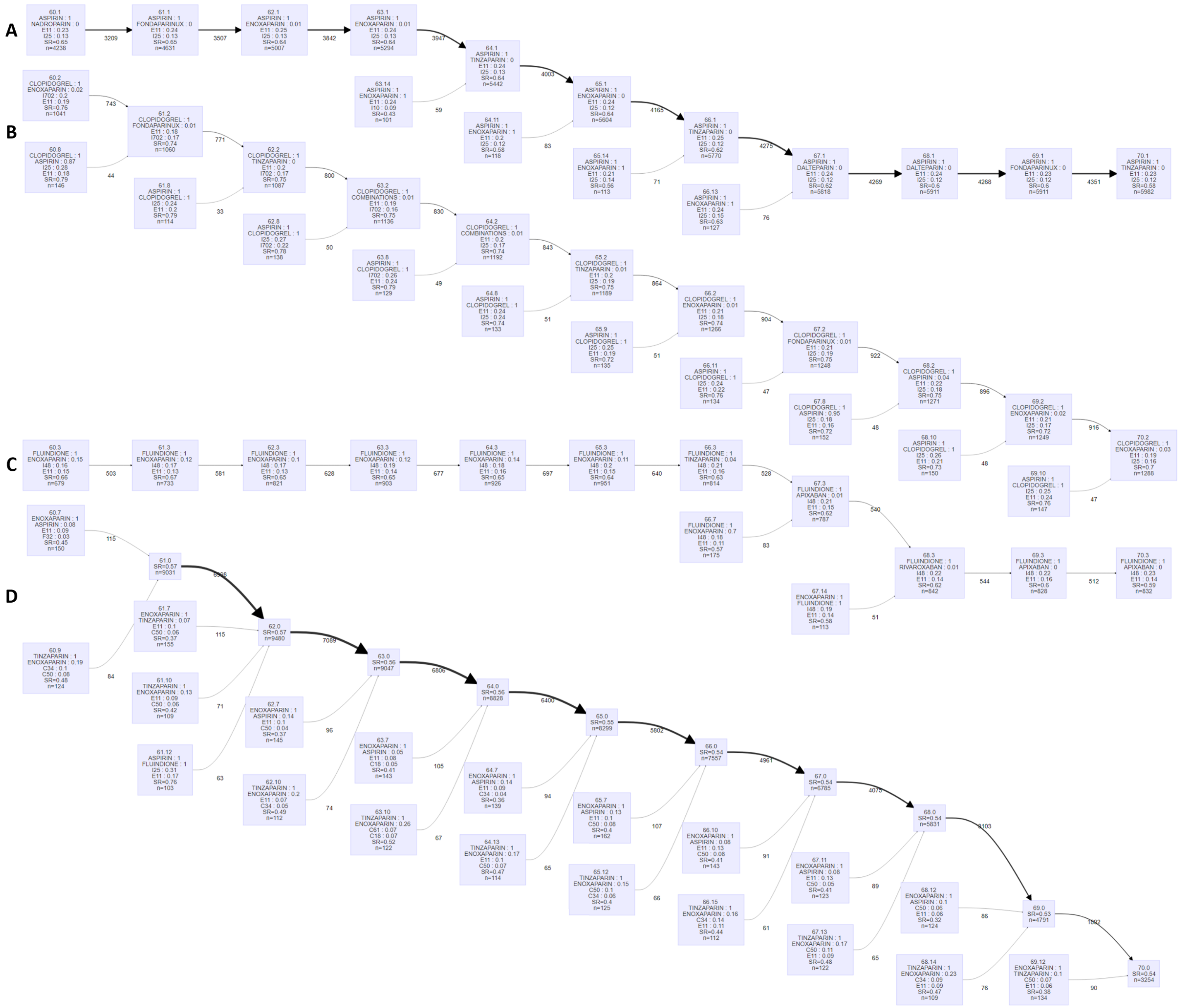


Figure S5: Network-based Cluster-trajectories part 1

Each block represents a cluster. Each cluster is named as follows: "x.y", with x the age time at which it was identified and y the number of the cluster. The clusters are characterized by the two most prescribed drugs (name, percentage of patients), the two most frequent long-term illnesses (ICD-10 code, percentage of patients), the sex ratio (SR) and the number of patients (n). The number under arrows is the number of patients in common between the two blocks. The arrow thickness is proportional to this number. Combinations: combinations of two platelet aggregation inhibitors.

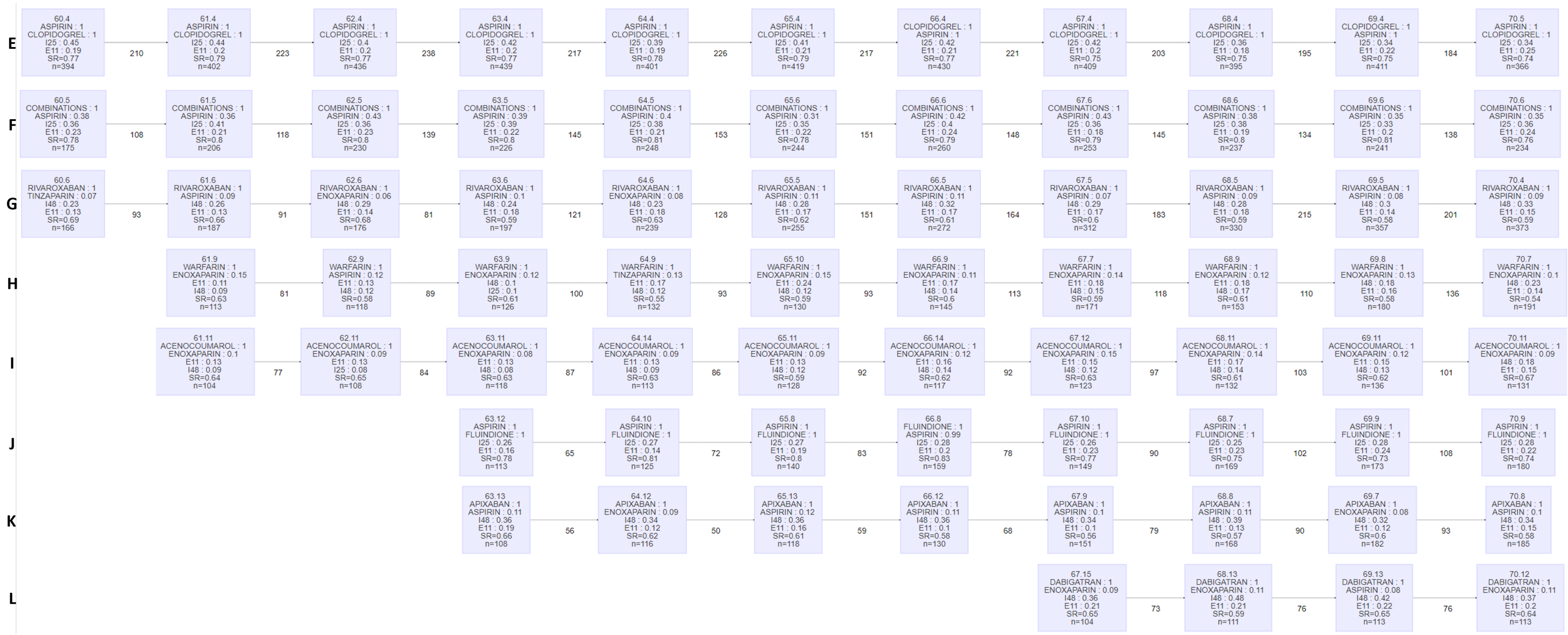

Figure S6: Network-based Cluster-trajectories part 2

Each block represents a cluster. Each cluster is named as follows: "x.y", with x the age time at which it was identified and y the number of the cluster. The clusters are characterized by the two most prescribed drugs (name, percentage of patients), the two most frequent long-term illnesses (ICD-10 code, percentage of patients), the sex ratio (SR) and the number of patients (n). The number under arrows is the number of patients in common between the two blocks. The arrow thickness is proportional to this number. Combinations: combinations of two platelet aggregation inhibitors.

#### S5 Assessing the optimal number of clusters using the silhouette score in the raw-data-based cluster-tracking approach

In the raw-data-based cluster-tracking approach, we applied a Kmeans to raw data, for each age considered. In Kmeans, the number of clusters must be specified *a priori*. We determined the optimal number of clusters per age by calculating the silhouette score (*Figure S5*).

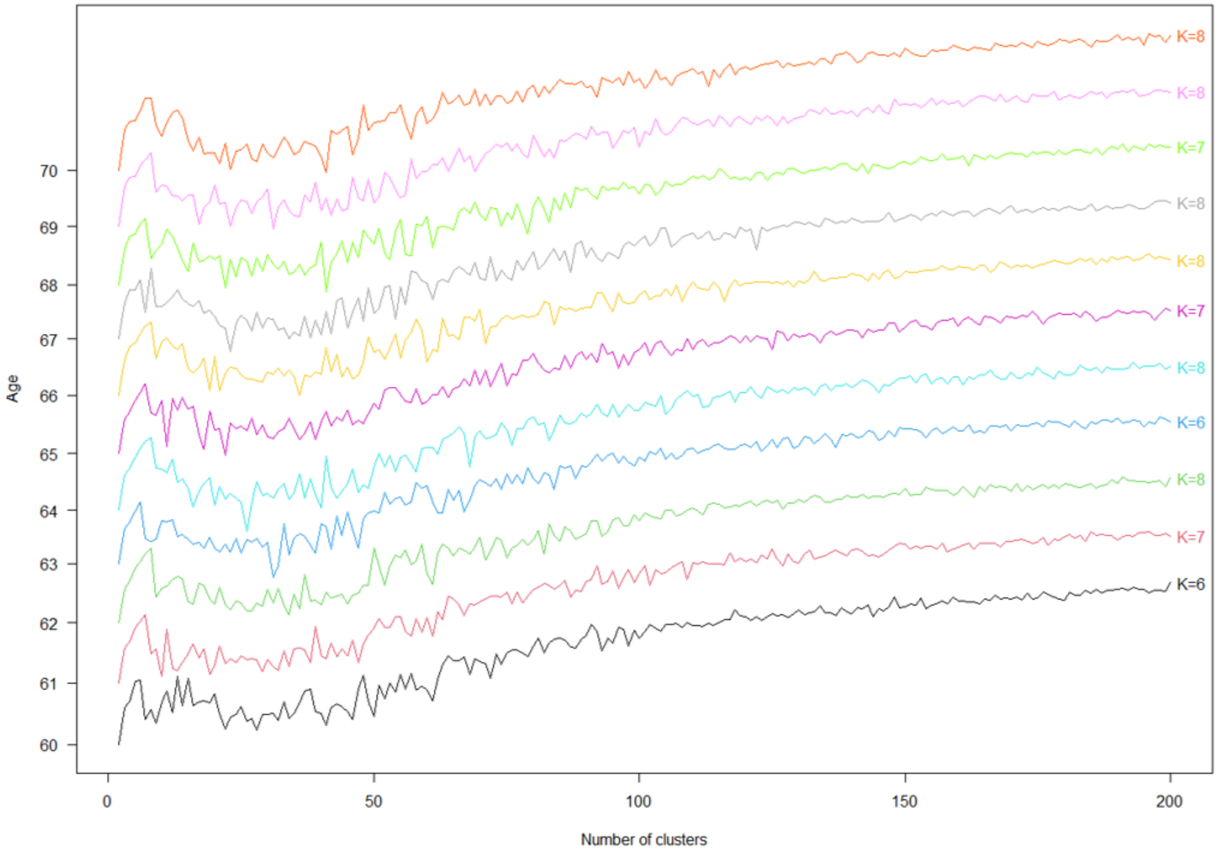


Figure S7: Silhouette score used to determine the optimal number of clusters at each age in the raw-data-based cluster-tracking approach

We calculated the silhouette score at each age, from 60 to 70 years old. We varied the number of clusters from 2 to 200. A specific optimal number of clusters $K$ is identified at each age.

#### S6 Cluster-trajectories identified with the raw-data-based cluster-tracking approach

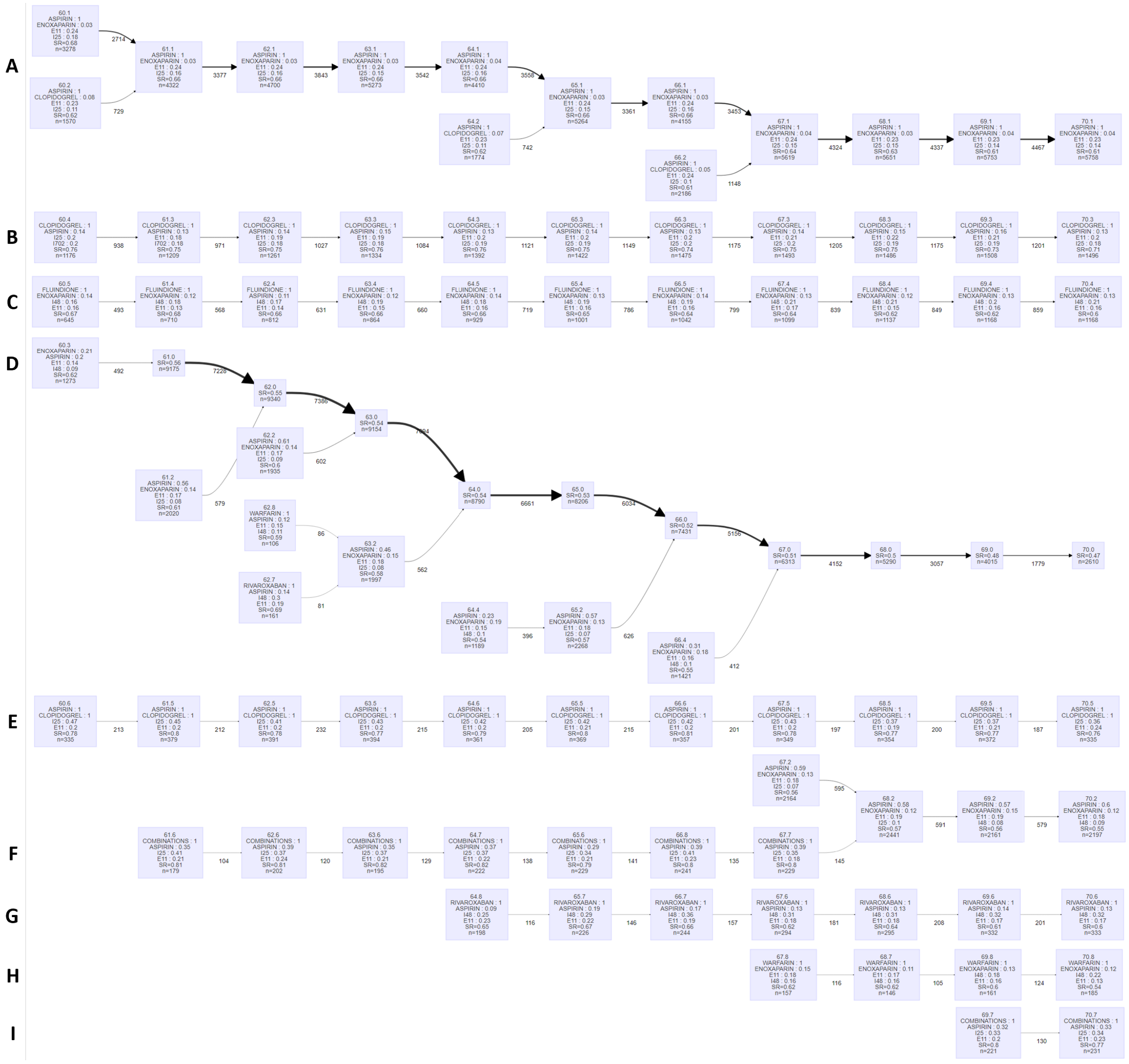


Figure S8: Cluster-trajectories in the raw-data-based cluster-tracking approach

Each block represents a cluster. Each cluster is named as follows: "x.y", with x the age time at which it was identified and y the number of the cluster. The clusters are characterized by the two most prescribed drugs (name, percentage of patients), the two most frequent long-term illnesses (ICD-10 code, percentage of patients), the sex ratio (SR) and the number of patients (n). The number under arrows is the number of patients in common between the two blocks. The arrow thickness is proportional to this number. Combinations: combinations of two platelet aggregation inhibitors.
